# B cell receptor analysis using single cell sequencing reveals preferential antibody expression in a HIV candidate vaccine study

**DOI:** 10.1101/2023.04.18.23288724

**Authors:** Irene Bassano, Simon Watson, Cinzia Malangone, Christopher L. Pinder, Kostas Paschos, Hannah M. Cheeseman, Robin J. Shattock, Paul Kellam

**Affiliations:** Imperial College London, Department of Infectious Disease, London W21PG, United Kingdom; Kymab, Babraham Research Campus, Cambridge CB22 3AT, United Kingdom; Illumina, Illumina Centre, 19 Granta Park, Great Abington, Cambridge, CB21 6DF, United Kingdom

**Keywords:** single cell RNA sequencing, B cells, V(D)J recombination, 10x Chromium, cell hashing

## Abstract

HIV (human immunodeficiency virus) is a virus that infects different cell types in the immune system. While different monoclonal antibodies have been identified, because of the antigenic diversity of HIV, defining broad neutralising antibodies (bnAbs) it has been very challenging. Using the 10x Chromium technology partitioning system, we performed single-cell sequencing of the B cell repertoire and analysed a cohort of 48 HIV-negative subjects randomised into 4 groups each receiving a different HIV vaccine regime to investigate the type of antibody response to each of the proteins in the vaccine groups, namely ConM SOSIP, EDC ConM SOSIP, ConS UFO and EDC ConS UFO. We recovered over >300,000 single cells and reconstructed full-length antibody heavy- and light-chain variable regions from individual memory B cells, plasmablasts, naïve B cells and activated B cells as well as retrieved their transcriptome profile to correctly separate the different populations within each individual. We identified seven clonotypes representing functional antibodies against vaccine candidate proteins. We reconstructed their phylogenetic tree and confirmed they were novel antibodies in response to HIV. These were mainly antibodies in response to two of the four protein candidates, namely ConS UFO and EDC ConS UFO.

## Introduction

B cells of the adaptive immune system produce antibodies that are specific for an antigen, with antibody diversity achieved by immunoglobulin gene V(D)J rearrangement and somatic hypermutation (SHM). During their development, immature B cells rearrange their heavy and light chain germline immunoglobulin genes to produce functional B cell receptors and exit the bone marrow as mature antigen naïve B cells, defined by expression of the cell surface markers CD19, CD20, IgM and IgD [1]. Upon antigen encounter, naïve B cells migrate to germinal centres (GCs) where they undergo class switch recombination (CSR) and antigen affinity maturation via extensive SHM [2]. B cells proliferate within GC follicles of secondary lymphoid organs, eventually differentiating into high affinity immunoglobulin-secreting plasma cells and memory B cells.

Memory B cells retain a “memory” of the antigen against which they matured, allowing for an anamnestic immune response upon subsequent exposures to an antigen. Pathogen genetic variation can limit the effectiveness of such an antibody response and in the case of persistent infection by HIV such variation leads to escape from antibody neutralisation. However, in some cases, HIV-positive patients develop broadly neutralising antibodies (bnAbs) [3], with high level of somatic hypermutation (SHM) and long CDR3s, which can recognize epitopes across a broad range of HIV genetic subtypes. This raises the possibility that if a vaccine can elicit such bnAbs then an effective HIV vaccine maybe achievable [4].

Indeed, over the years several HIV vaccine candidates have been proposed based on their efficacy, safety and immunogenicity: these antibodies target the native envelope protein (Env) and include recombinant Env proteins with or without adjuvants, HIV viral like particles (VLPs), HIV DNA plasmids, or replication-competent or incompetent viral vectors[5]. The main goal is for these antibodies to block either the Env-CD4 receptor or the virion fusion with host cells [6]. BnAbs are known to target the following six epitopes on the Env protein: MPER (membrane proximal external region) of gp41, V1V2-glycan, the outer domain glycan, V3-glycan and the CD4 binding site [7].

Current vaccine developments commonly face many obstacles, one of which being the high diversity of the circulating virus itself. Despite this, many trials have been developed, with different degrees of success. Among those in the early phase clinical vaccine trial, it is worth mentioning the IAV G001 (NCT03547245) study which used eOD-GT8 as immunogen (a self-assembling nanoparticle with HIV Env protein) and AS01B as adjuvant. This trial ran in USA in both women and men between June 2018 and December 2020. Another clinical trial, the EAVI2020_01 (NCT03816137) used ConM SOSIP, EDC ConM SOSIP, ConS UFO, EDC ConS UFO and mosaic SOSIPs and MPLA as adjuvant and ran in UK in both women and men between March 2019 to present. Among the HIV vaccine efficacy trials, VAX003 and 004 were one of the firft ones but did not prevent HIV infection in both men and women that were enrolled. On the other hand, the RV144 (NCT00223080) is one of the most successful to date, with a 31% efficacy. This trial employed a pox-protein prime-boost strategy with ALVAC-HIV (vCP1521) expressing Gag and Pro (subtype B LAI) CRF01_AE gp120 (92TH023) linked to transmembrane anchoring portion of gp41 (LAI) AIDSVAX B/E absorbed to Aluminium hydroxide. Another notable efficacy trial was led by Johnson and Johnson between November 2017 and July 2021 in south Africa. The Imbokodo HVTN 705 (NCT03060629) Phase 2b vaccine efficacy trial employed Ad26.Mos4.HIV Subtype C gp140 bound to aluminium phosphate. Current results were expected in July 2022 but the trial was halted following a 25.5% efficacy result, failing to offer substantial protection against HIV [8].

The concept of designing a vaccine starting with known antibodies is known as “reverse vaccinology” [9] and can also be based on the information retrieved from deep sequencing of the BCR repertoire, which allows us to determine the type of antibodies produced by an HIV infected person or patients under clinical trials. This is different from the classical methodology of retrieving antibody repertoire by sequencing antigen sorted cells [10]. In the past decade, RNA-Seq has been widely used to assess gene expression levels within cell populations. Comparison of treated and untreated samples has given us an exhaustive overview of how the transcriptome can change under different conditions providing an insight into what an infection, antigen, or cancerous status would trigger within the cells. In addition, the more sophisticated method of single cell RNA-Seq (sc-RNA-Seq) has been developed over the last decade. Thanks to techniques such as SMARTSeq and single cell droplets methods, we are now able to sequence thousands of cells simultaneously and discover the diversity of subpopulations which otherwise would be lost in bulk sequencing. In addition, with the advent of CiteSeq and hashing methodologies, we are now for the first time able to simultaneously assess different samples in single encapsulation runs saving both costs and time towards the final libraries. These methods have been successfully used to study both B and T cell repertoires.

Here we applied single cell sequencing to B cell populations to investigate the dynamics of antibody diversity following vaccination with novel HIV immunogens. We show that coupled B cell receptor and transcriptome sequencing allows simultaneous B cell phenotyping and BCR repertoire analysis, and that B cell lineages in peripheral blood B cells can converge in different participants following vaccination.

## Material and methods

### Clinical trial patients

Samples from day 7 following the first vaccine dose for 48 HIV-negative participants of the EAVI2020_01 (NCT03816137) clinical trial against HIV [11] designed to assess if different prime-boost combinations with novel HIV immunogens could influence serum neutralising antibody breadth, were used here. These 48 patients were randomised into 5 groups each receiving a different vaccine regime (Supplementary Table 1). As two groups (C and E) had the same vaccine regime for first injection and different vaccine regimes for the boosts, for this initial time point we combined these two groups. Here therefore, we have used samples from participants immunised with A) 100µg of an HIV trimer based on a consensus sequence of all HIV-1 group M isolates from 2004 (ConM SOSIP), with (Group B) or without (Group A) EDC (1-ethyl-3-(3-dimethylaminopropyl) carbodiimide) (EDC ConM SOSIP), or Group D, which received 100µg of ConS UFO (Uncleaved pre-fusion optimised) HIV trimer, based on a consensus sequence from 2001, with (Group D) or without (Group C and E) addition of EDC (EDC ConS UFO) [12-16] (Supplementary Table1). The two vaccine protein candidates represent stabilised versions of the native Env protein. Briefly, the ConM SOSIP contains an extra intramolecular disulphide bond (SOS) to stabilize the gp120/gp41 interaction, in addition to a single amino acid point mutation at position 559 where an Isoleucine is replaced by a Proline (IP) [17-19]. ConS UFO forms a closed pre-fusion membrane-bound Env trimer and contains a short flexible amino acid linker to join gp120 and gp41 subunits. Therefore, ConM and ConS use two different approaches to prevent Env dissociation into monomers [13], while the addition of EDC, a crosslinking agent, preserves the nature of the bnAbs.

### Isolation of B cell populations

Up to 3x10^7^ frozen peripheral blood mononuclear cells (PBMCs) from 48 HIV-negative donors were collected 7 days post vaccination. The immediate response to the vaccine will be led by antibody-secreting plasmablasts, while 7 days later memory B cells will proliferate with the membrane-bound form of the antibodies.

Memory B cell and plasmablasts were isolated using the Miltenyi Pan B and CD27 Microbead isolation kits according to manufacturer’s instructions. Briefly, frozen PBMC were washed in RPMI media containing 10% FBS and twice with PBS to remove excess DMSO. Cells were subsequently incubated with a Pan-B cell biotin-antibody cocktail to remove non-B cells by negative selection. Cell populations from the eluted fraction were then enriched by positive selection using CD27 microbeads to capture all cells expressing CD27 on their surface. Although the two kits were specific for memory B cells and plasmablasts from the same pool, we were also able to simultaneously isolate naïve and activated B cells based on the expression of certain cell marker transcripts during bioinformatic analysis. Enriched cell populations were then counted and assessed for viability. Only cell populations with a viability above 85-90% were considered for single cell encapsulation.

### Cell hashing and 10x microencapsulation

Purified B cell populations underwent the cell hashing protocol as described in [20]. The following hashing antibodies from Biolegend were used at a final concentration of 1ug/ml: TotalSeqC-251, 252, 253, 254, 255, 256, 257, 258, 258, 259, and 260. All of the TotalSeq –C antibodies use a combination of two clones that recognize CD298 and β2 microglobulin, respectively. Following hashing of the single donors, cell numbers and viabilities were determined, samples pooled, and then loaded onto a 10x Chromium at 17000 cells per lane, with an expected recovery of 10000 cells. Libraries were prepared as per protocol [21]. Three types of libraries were prepared: V(D)J, whole transcriptome (GEX, gene expression) and feature barcoding (FB). The latter was used to deconvolve to each individual sample based on the unique sample barcode present. Libraries were normalised to 4nM and sequenced on Illumina platforms.

### Sequencing and data analysis

Libraries were sequenced using an HiSeq 4000 or Novaseq6 from Novogene. Fastq samples were processed for quality control, adapters removed, and sequence analysis performed using CellRanger v3.1 and v5. Hashed samples were demultiplexed using CiteSeq v1.4.0 together with Seurat v.3.0 and further partitioned using Loupe Cell Browser v4.0.0 (10x Genomics) based on the sequence tag of the cell hashing antibodies. Duplets, which represent single droplets containing more than one encapsulated cell, were identified either using Scrublet [22], CiteSeq, Seurat or Loupe Cell Browser. V(D)J sequences were assembled from cells that expressed both paired heavy and light chain mRNA. Of these, only full length and productive IgH and IgL transcripts were considered for final analysis (Figure 1) using enclone (https://10xgenomics.github.io/enclone/). Cell type separation was performed using a list of transcription factors which define B cell subpopulations, specifically for memory B cells, plasmablasts, activated B cells and naïve B cells (ref) (Supplementary Table 2), thereby allowing IgH and IgL sequences and their respective clonotypes to be assigned using gene expression data to B cell type gene expression using Loupe Cell Browser. Additional custom scripts were used to calculate V, D, J, and C gene usage and generate tables with V(D)J gene combinations for display using Circos plots.

**Figure 1.**
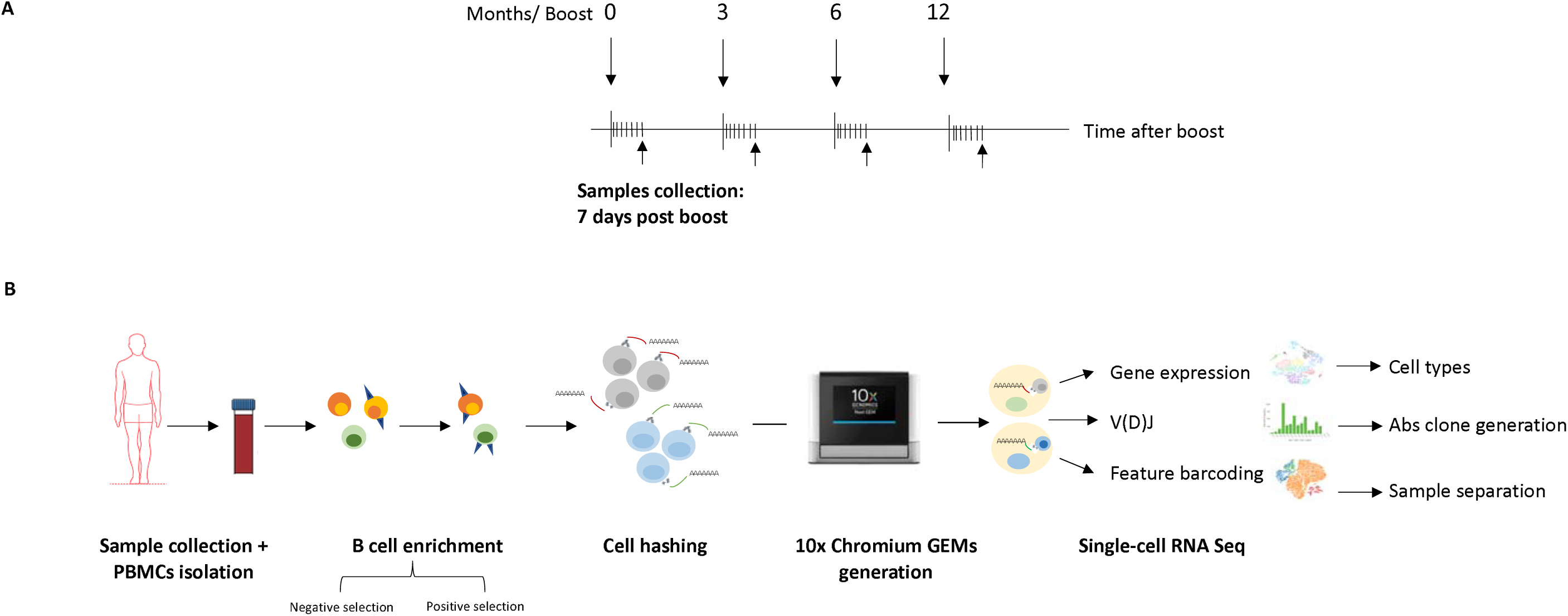
Schematic overview describing the process from sample collection to data analysis. A. Timeline of the vaccine program showing which samples were used for this work. B. Figure describing the steps from blood collection to encapsulation up to sequencing and data analysis.

### Phylogenetic analysis

Nucleic acid sequences of the V_H_ and V_L_ domain of each antibody in a clonogroup were concatenated and exported in FASTA format. The inferred germline IGHV, IGHJ, IGLV, and IGLJ genes were then added to the FASTA file. IGHD genes were not included due to the difficulty in unambiguously inferring the correct gene. The germline sequence was manually aligned to the rest of the concatenated V_H_/V_L_ sequences from the clones in the clonogroup using AliView version 1.26 [23]. Neighbour-joining (NJ) tree was inferred from the alignment using the phangorn R library and the resultant tree was rooted on the germline taxon. Metadata such as V_H_ isotype, cell type, sample group and sample ID were associated to the phylogenetic tree using the ggtree R library.

## Results

### Cell recovery and sample separation using expression profile of the hashing antibodies

Human peripheral blood B cells consist mainly of naïve and memory B cells, however, immediately following antigen encounter a transient population of plasmablast B cells can be found in the peripheral blood corresponding to B cell affinity maturation in secondary lymphoid organs. Therefore, by obtaining PBMC samples on day 7 following immunisation with the different HIV immunogens we can maximise the chance of detecting vaccine antigen-responsive plasmablasts and then relate these to the subset of recently produced memory B cells to the same immunogen. We therefore enriched for plasmablasts and memory B cells from the peripheral blood at this timepoint using magnetic cell sorting for CD27 positive B cells. To simultaneously obtain B cell receptor sequences and cell type identification, we performed single cell sequencing by combining V(D)J sequencing and whole transcriptome sequencing. Furthermore, we were able to pool multiple patient samples by labelling them with hashing antibodies linked to a unique sample identifier sequence tag for each sample. Using these methods, we were able to combine up to 4 different patient samples in one 10x run (Figure 1b). Cell hashing of samples allows an estimation of the number of times ‘single cell’ sequencing occurs with a droplet containing more than one cell. Such ‘duplets’ were identified where an apparent single cell transcriptome contained two distinct cell-hashing sequences, rather than the assumed single hashing antibody sequence tag present per cell. For each multiplex hashed sample, we pairwise compared hashing sequence where single cells that were not only on the x or y axis represent potential duplets (Supplementary Figure 1a). Duplets were also identified using Seurat/CiteSeq (including for non-hashed, single sample single cell sequences) and by using Single-Cell Remover of Doublets (Scrublet)(25). Analysis of tSNE gene expression showed such duplets were not clustered in a distinct gene expression space for B cells and so could not be triaged based on a gene expression pattern (Supplementary Figure 1b). Combining these multiple duplet removal methods yielded a final single cell dataset for further analysis.

### B cell populations and transcriptional profile in ConM SOSIP and ConS UFO samples

Transcription factor gene expression can be used to define cell types and therefore we compiled a list of B cell transcription factors [24, 25] for the classification of B cell types from the scRNAseq data. (Supplementary Table 1). Four B cell populations were defined: memory B cells (MB), plasmablasts (PB), activated B cells (AC) and naïve B cells (N), along with gene expression markers for Leukocytes, T cells, erythrocytes, and monocytes. Cells that did not fall into these criteria were classified as undefined B cells (UB) (Figure 2b and 3a,b). These latter cells had a very low level of UMIs for the transcription factors we used to differentiate into the population of interest, but nevertheless were grouped into one cluster and analysed with the other cell types. In addition, we added to the list T cell and monocyte markers to remove any possible contamination from the magnetic separation of the B cell populations. The clustering method showed that although some cells did express transcription factors found in T cells and monocytes, it also highlighted that they had higher expression of the markers used for B cells. Consistent with the B cell enrichment methods for memory B cells and plasmablasts, all vaccine groups showed an abundance of cells that could be defined as either memory B cells or plasmablasts by transcription factor expression (Figure 2a, b). 39% of the total single cells consisted of MB, with 7% of total cell being classified as PB. 1% of cells were classified as activated B cells and 1% as Naïve B cells, perhaps reflecting low level contamination of other B cells during the cell enrichment protocol consistent with FACS sorting data [25]. By combining all sample data, it is clear that these classifications were present in all samples and vaccine groups. Due to multiple duplet identification methods, we conservatively assigned 47% of cells as duplets. Through combined V(D)J seq we were able to assemble complete paired IgH and IgL sequences from the majority of single cells. If paired IgH&L was not detected these were assigned as V(D)J negative cells. Interestingly most V(D)J negative cells were assigned as duplets, suggesting that as expected a duplet transcriptome would render impossible the assembly of paired IgH&L sequences.

**Figure 2.**
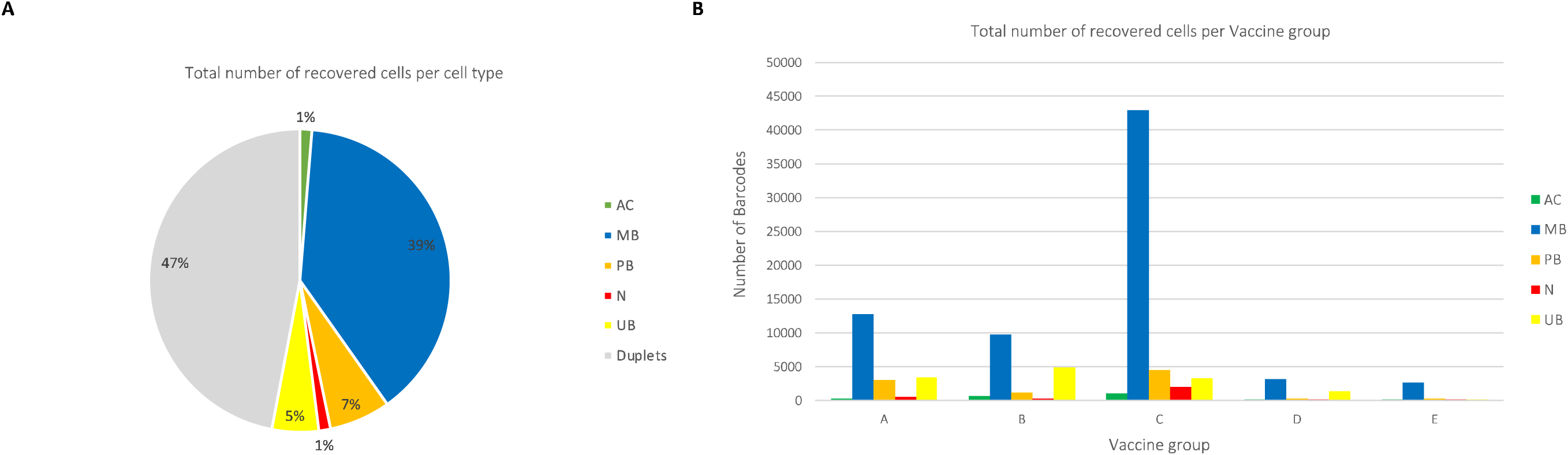
Number of recovered cell types in all the samples removed duplets and V(D)J negative cells, we recovered ∼1 (A) and within each of the vaccine groups (B). On average, once we 150000 total cells.

**Figure 3A-B.**
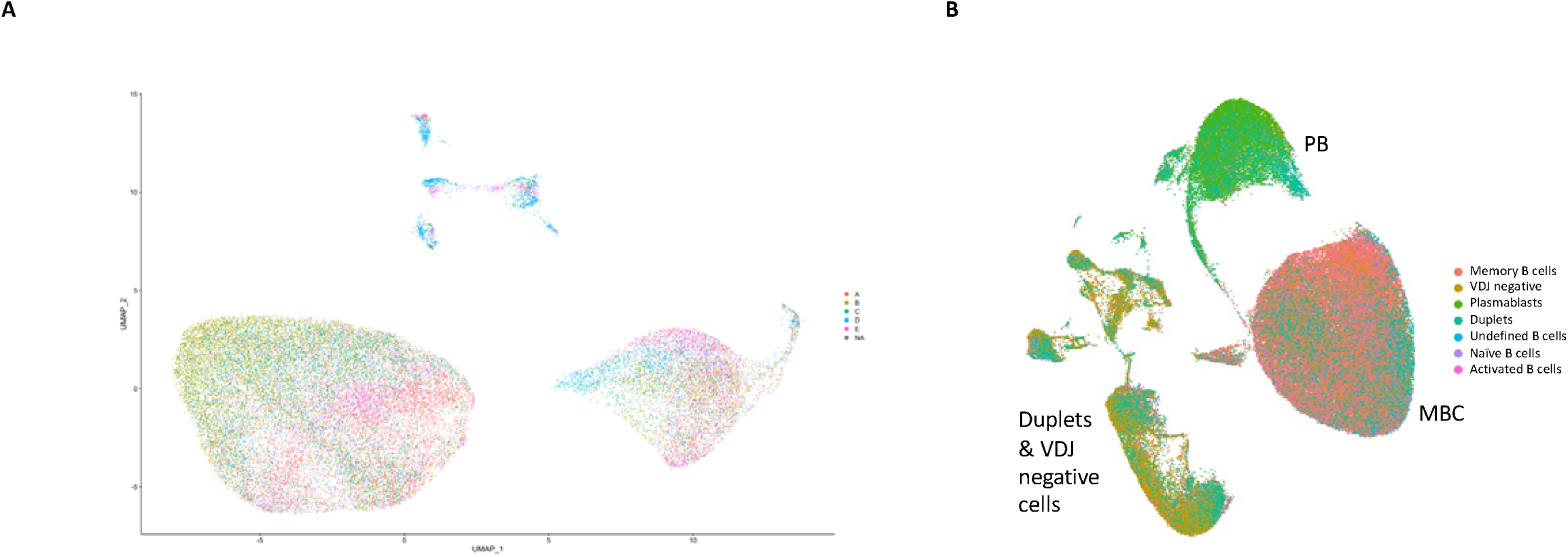
Generation of cell clusters. A. All sam les were grouped together following assignment of of the vaccine group they belonged to. NA represent a group of cells which were either duplets and/or V(D)J negative. B. Cell type separation based on internal transcrip ional markers. The figure shows the different B cell subpopulation retrieved after separating duplets and V(D)J negative cells. Samples that showed expression of barcodes which didn’t fall in any of the B cell populations we retrieved based on the transcriptional markers used were defined as undefined B cells (UB). As shown in the figure most of the population is represented by memory B cells, followed by plasmablasts, activat d B cells and naïve B cells (excluding duplet barcodes).

### V(D)J gene usage and CDR3 region frequency

We determined the IgH and IgL gene usage irrespective of B cell type, for productive (open reading frame), full length, paired heavy and light chain containing cells (Figure 4a) per immunisation group. For all immunisation groups, IGHV3-23, IGHD3-22, IGHJ4 and the IGHM isotype were the most common. Interestingly, IGH genes IGHV3-53 and IGHV3-66 commonly part of Class 1 nAbs were also commonly detected. For the lambda light chain (Figure 4b), IGLV2-14, IGLV1-44 and IGLV1-40 were the most common V segment among the groups. For the kappa light chain, IGKV3-20 was the most used segment for vaccine groups A, B and D, while IGKV4-1 was the most used in group C. For the J gene segment in the kappa light chains, we found that all the groups predominantly used the IGKJ1 gene segment (Figure 4b). All groups preferentially use the kappa light chain (Figure 4d). Overall, IgHM, G2, A1 are the most common isotypes across all groups (Figure 4c). We determined the most common CDR3 sequences within each immunisation group. As shown in Figure 4e, each group had a preferential CDR3 sequence: those with the highest frequency were CARASGYYPYYLDYW in group A, CASRSGRNYYGMDVW in group B, CAKWRRRDGYNFESW in group C and D (Figure 4E). IGH and IGL V(D)J recombination maps were visualised using Circos for all the 48 subjects within each vaccine group (Figure 5a) and for each individual group (Figure 5a,b) but split across the cell types of interest (MB and PB) for the most frequent clonotypes (top 75). Whilst qualitative in analysis, it is clear that all patient groups and MB and PB B cell types have considerable immunoglobin gene recombination diversity.

**Figure 4A.**
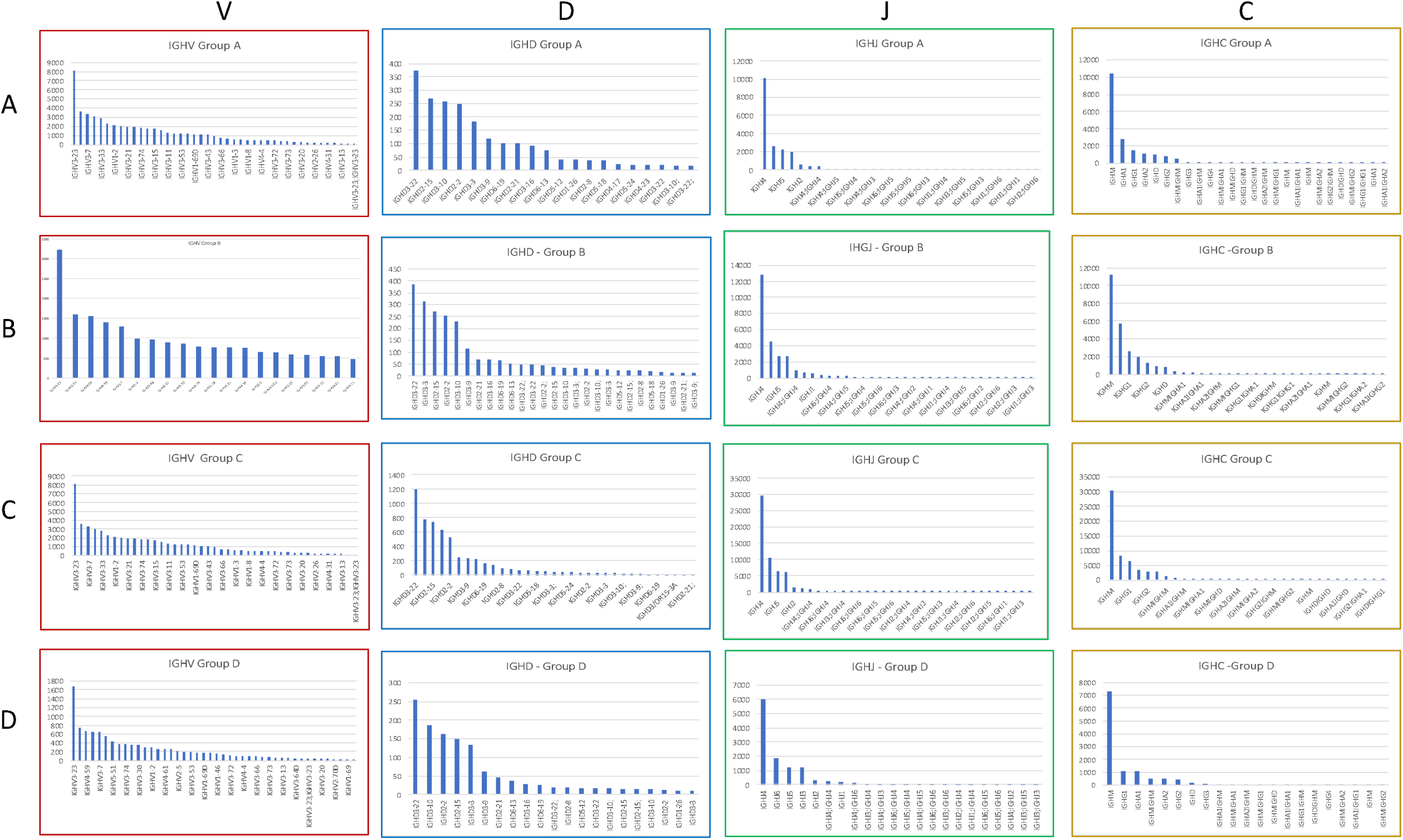
Plots of heavy chain V, D, J, C gene usage among the 4 vaccine groups. Only the to highly expressed genes were chosen for plotting. Columns represent the four ge es and rows the four vaccine groups for which we calculated gene levels expression. For all groups, among the V gene, IGHV3-23 was the most used; for the D gene, IGHD3-22 was common to all groups; for the J gene, IGHJ4 was exp essed highly in all groups and the most constant gene was IGHM.

**Figure 4B.**
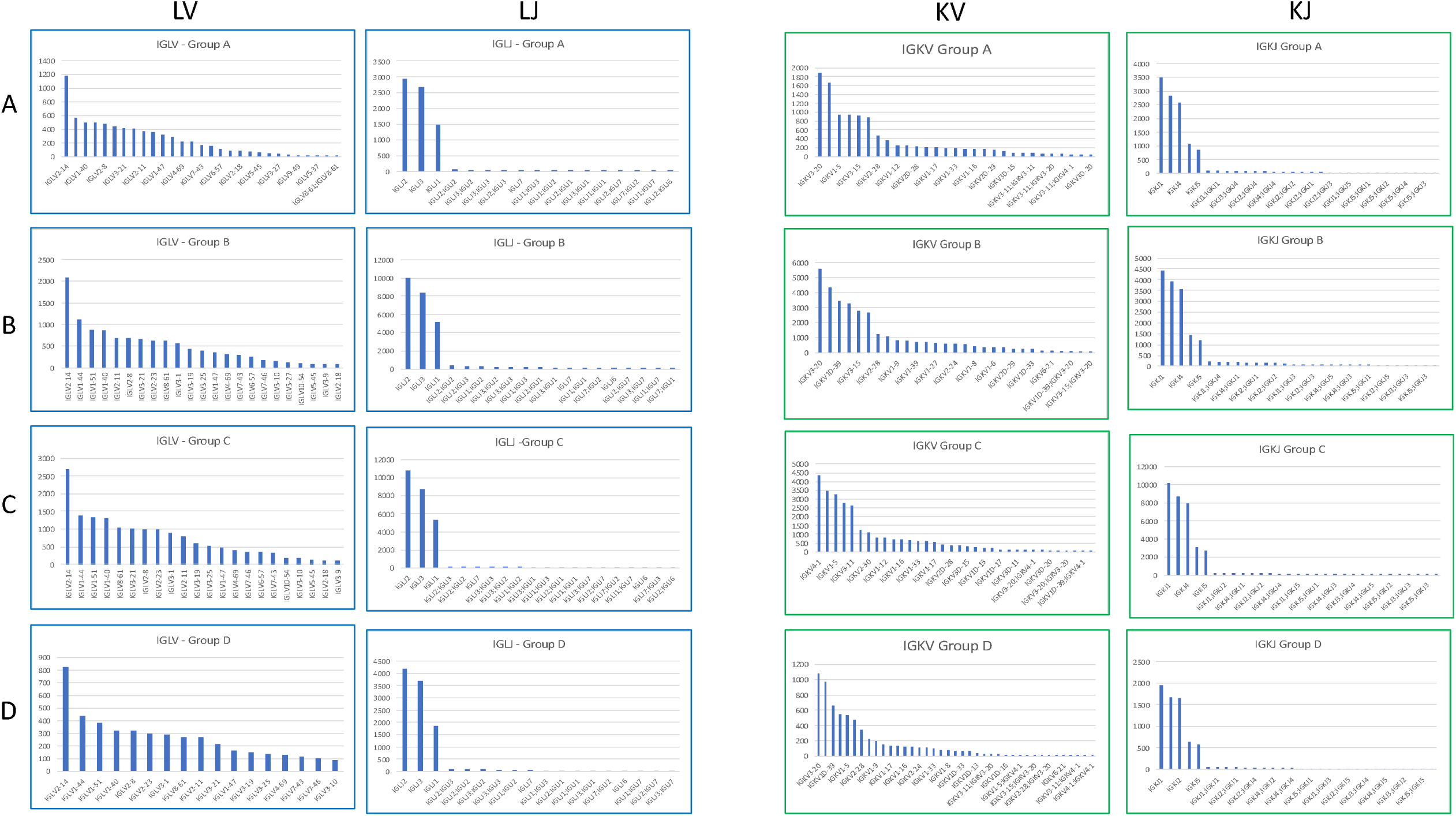
Plots of light chain V, J, C gene usage among the 4 vaccine groups. Left side for the Lambda light chain and right side for the Kappa light chain. The figure shows that IGLV2-14 and IGLJ2 were the top highly expressed genes for the Lambda light chain, while for the Kappa light chain IGKV3-20 was highly expressed in groups A, B, and D. Group C had IGKV4-1 as the top used gene. For the Kappa J gene, IGKJ1 was highly expressed in all four groups.

**Figure 4C.**
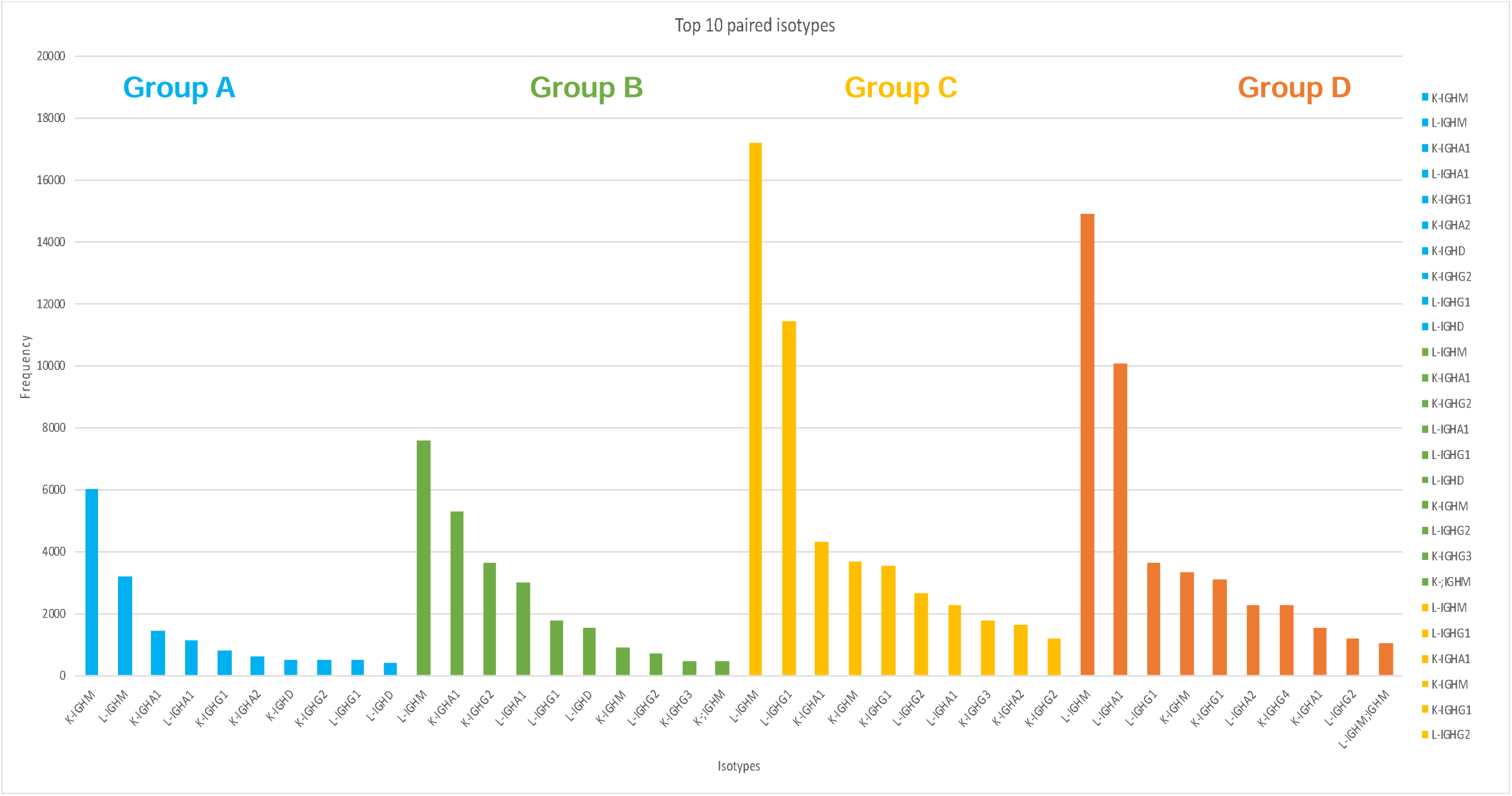
Frequency plots of top paired isotypes per vaccine group. The figure shows the preferential used of the paired isotypes for each vaccine group represented in four different colors: group A, blue, group B, green, group C, yellow and group D, orange. The following pairs were on top of each group: IGHM-K for group A, IG M-L for group B, IGHG1-L for group C and IGHG1-L for group D.

**Figure 4D.**
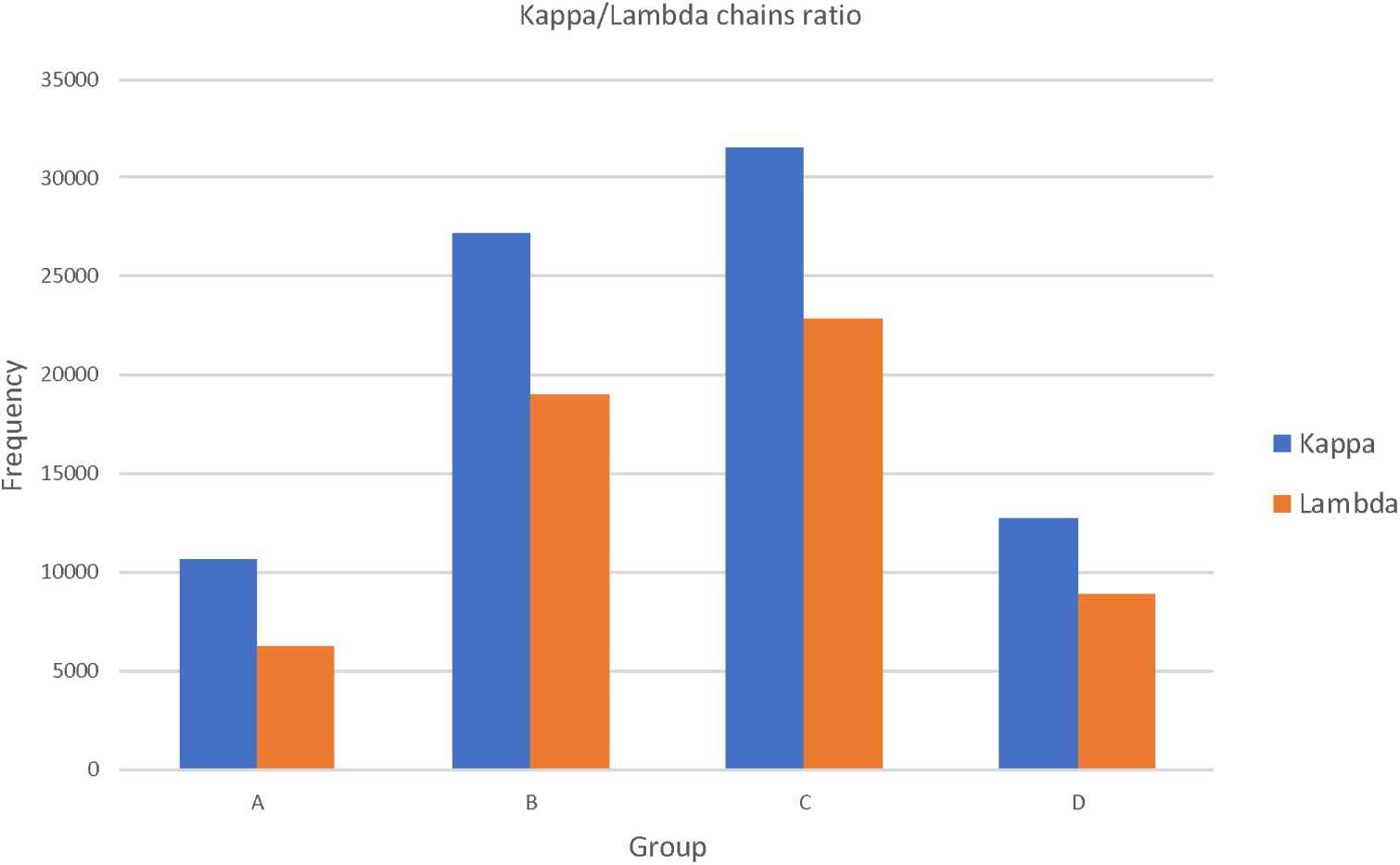
Frequency plot showing ratio of Kappa vs Lambda usage in each vaccine group. The figure shows the ration of Kappa vs Lambda light chain usage in each group. The frequency is higher for Kappa light chain in all groups compared t the Lambda chain.

**Figure 4E.**
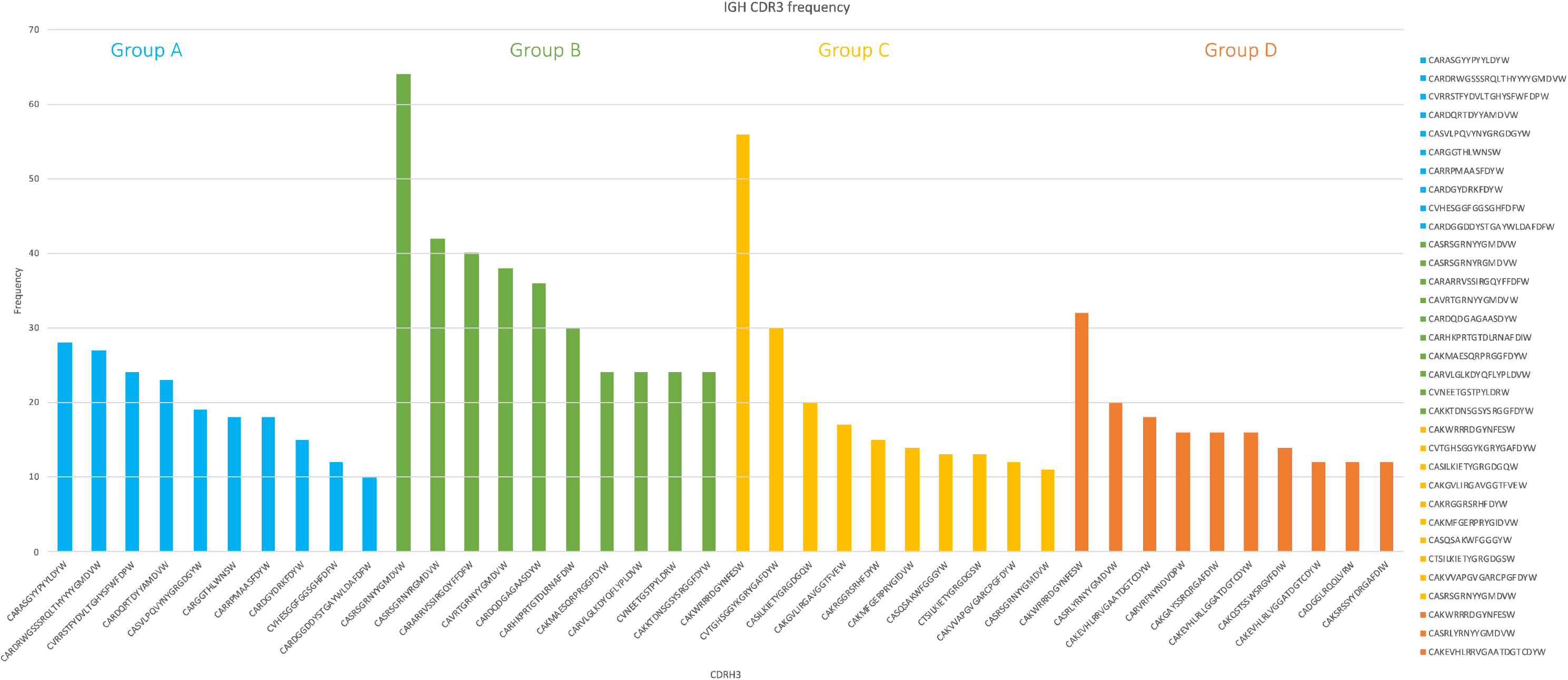
Frequency plot of the top 10 heavy CDR3 regions. The figure shows the top 10 CDR3 most used regions among the four vaccine groups represented in four different colors: group A, blue, group B, gr en, group C, yellow and group D, orange. Groups B and D share the same CDR3 region in their top usage compared to group A and C (see text for more detail about the CDR3 region sequence).

**Figure 5A.**
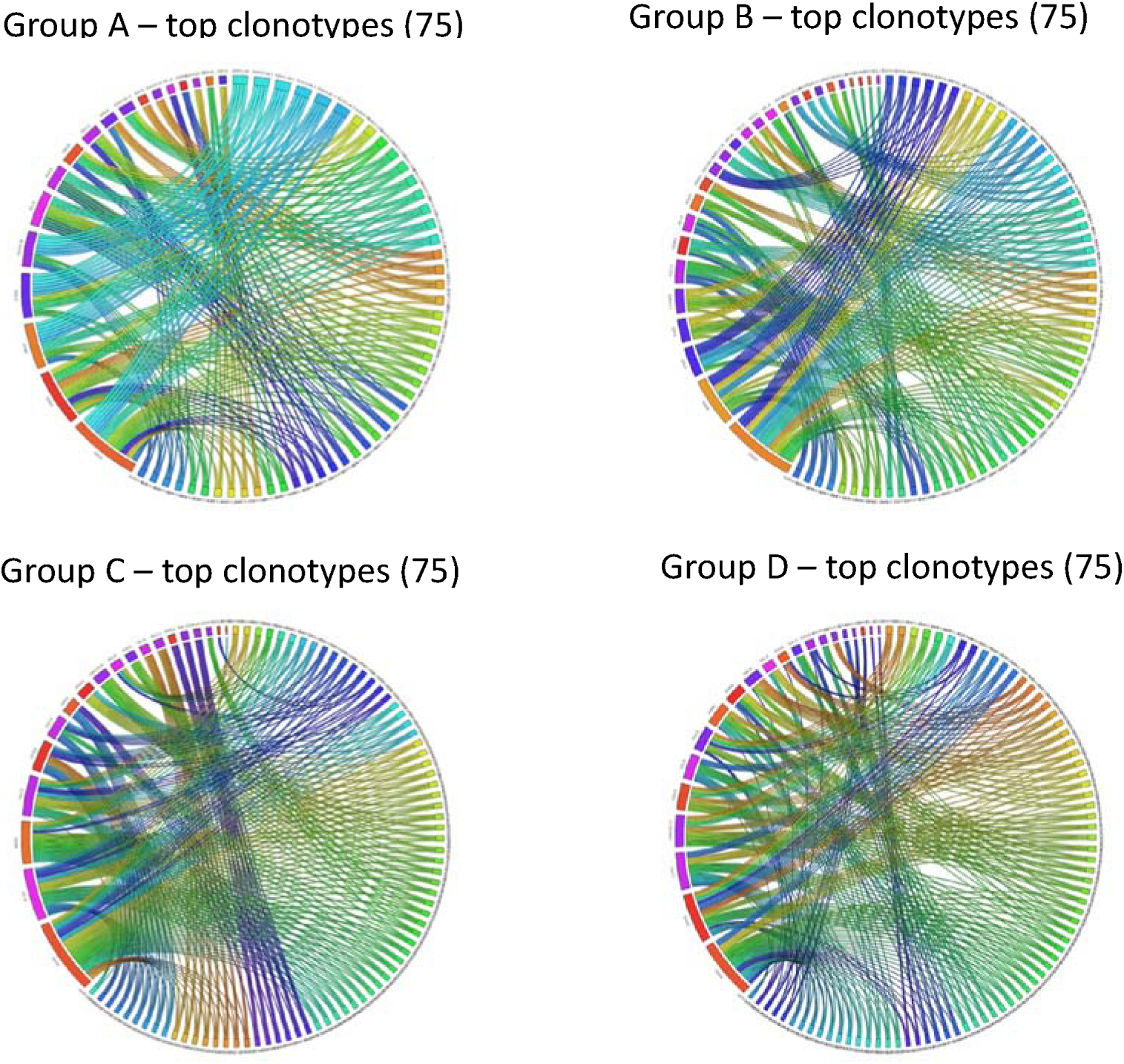
Circos plot showing the highest recombining events as clonotypes for the four vaccine groups. The figure shows only the top 75 recombination events. Given the high number of genes used (see text file in supplementary to read the gene names), the circos plot is only used as a visual representation of the clonotypes. All together the figure shows that there isn’t a particular clonotype with a high frequency of recombination (width of curved lines), instead these events are evenly distributed among several clonotypes.

**Figure 5B.**
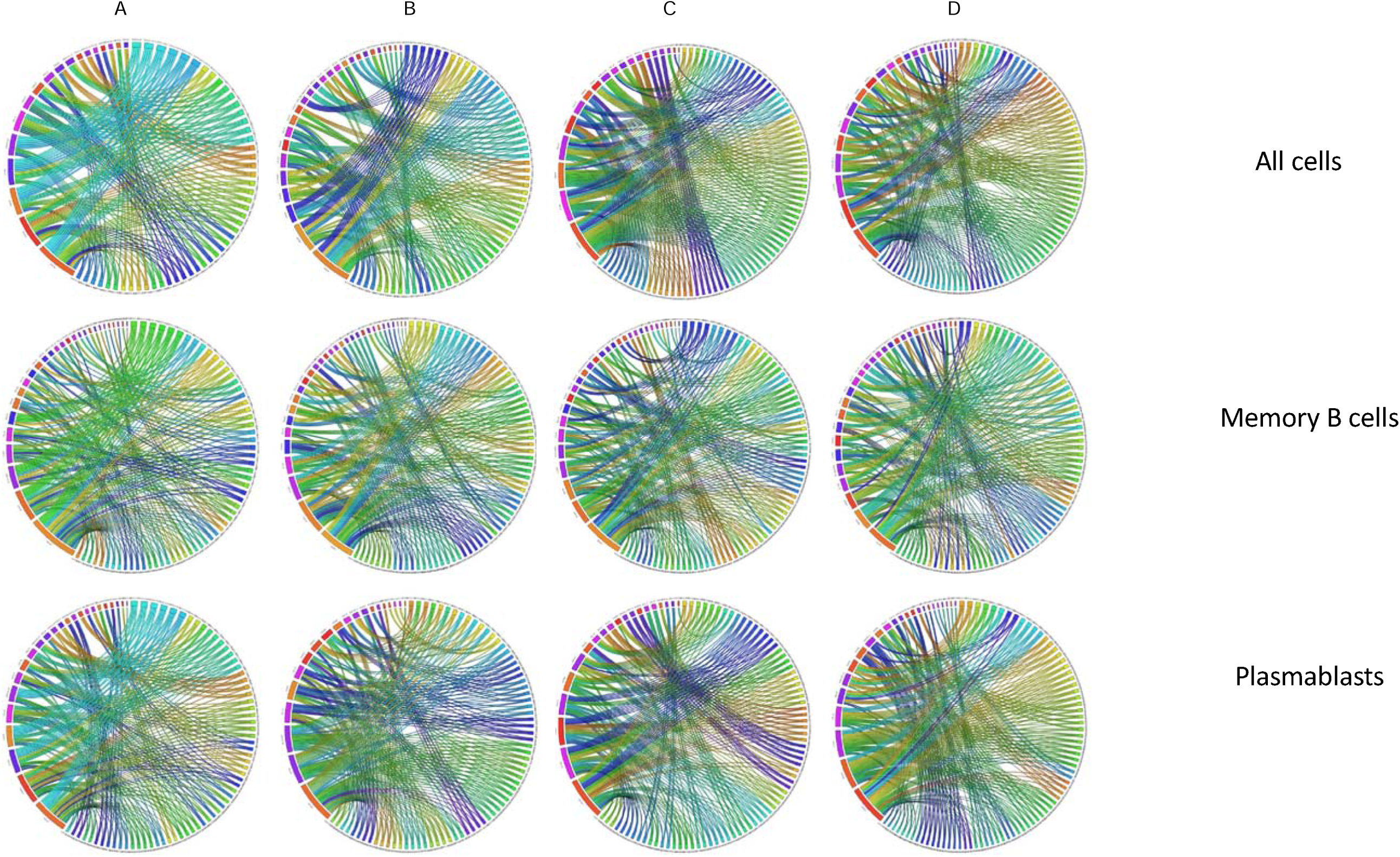
Circos plot of clonotypes distributions divi ed by cell types. Circos plot were generate specifically highlighting all cell types (top row) or focusing only onto memory B cells (middle row) and plasmablasts cells (bottom row). As shown in Figure 5A, there isn’t a particular clonotype with a high frequency of recombination (are evenly distributed among several clonotypes.

### V(D)J clonotype analysis and isotype usage between groups and cell types

By combining B cell receptor repertoires with B-cell identity, it is possible to define more precisely antigen experienced B-cells. To do this we identified BCR clonotypes (Fire 6A) from IgH&L paired single cell sequencing, assigned them their immunoglobulin isotype (Fig 6B) and assigned, by transcription factor expression, the B cell phenotype (Fig 6C). Given that peripheral blood B cells were isolated 7 days after antigen exposure, we defined antigen experience cells as those of an expanded clonotype, where the immunoglobulin isotype was switched IgG, IgA or IgE and where the cells of the clonotype were ABCs, or MBC combined with PBs. Clonotypes were used as the organising feature and these multi-level B cell clonotypes were visualised using Enclone honeycomb plots (Figure 6A). Here we show all the clones from each patient (one colour per patient) plotted onto one honeycomb plot to visualize possible shared clonotypes (represented by mixed colours combs). We represented this also by isotypes (one colour per isotype) as well as cell types (one colour or cell type). A small number of clonotypes had a single antibody isotype, namely 24 IgM clonotypes and 13 IgD clonotypes. Similarly, the majority clonotypes were made up only of plasmablasts, or contained memory B cells combined with other B cell types, especially plasmablasts.

**Figure 6A.**
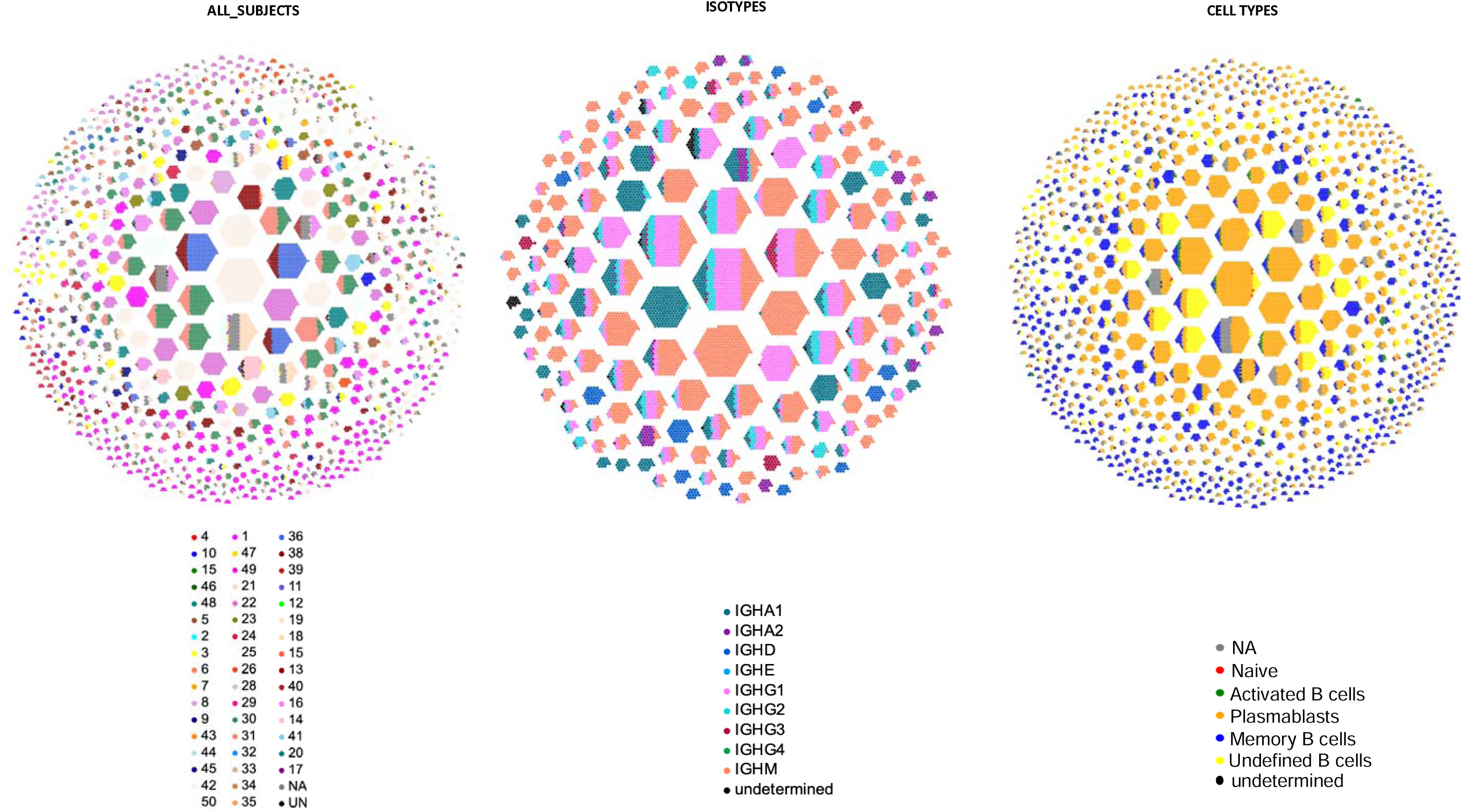
Honeycomb plots of shared clonotypes. The figures shows the expression of clonotype by taking all subjects (left), highlighting the expression of isotypes (middle) or cell types (right). Colors indicate either the subjects from all the 4 vaccine groups, the type of isotypes used or the type cells. Take al together, each clonotype is shown in the same shape in each plot. Each comb where more than one color is shared, represents, e.g., a subject with more than one isotypes across different cell types. Sometimes, a clonotype is shared among two or one color, e.g. more than one subject).

**Figure 6B.**
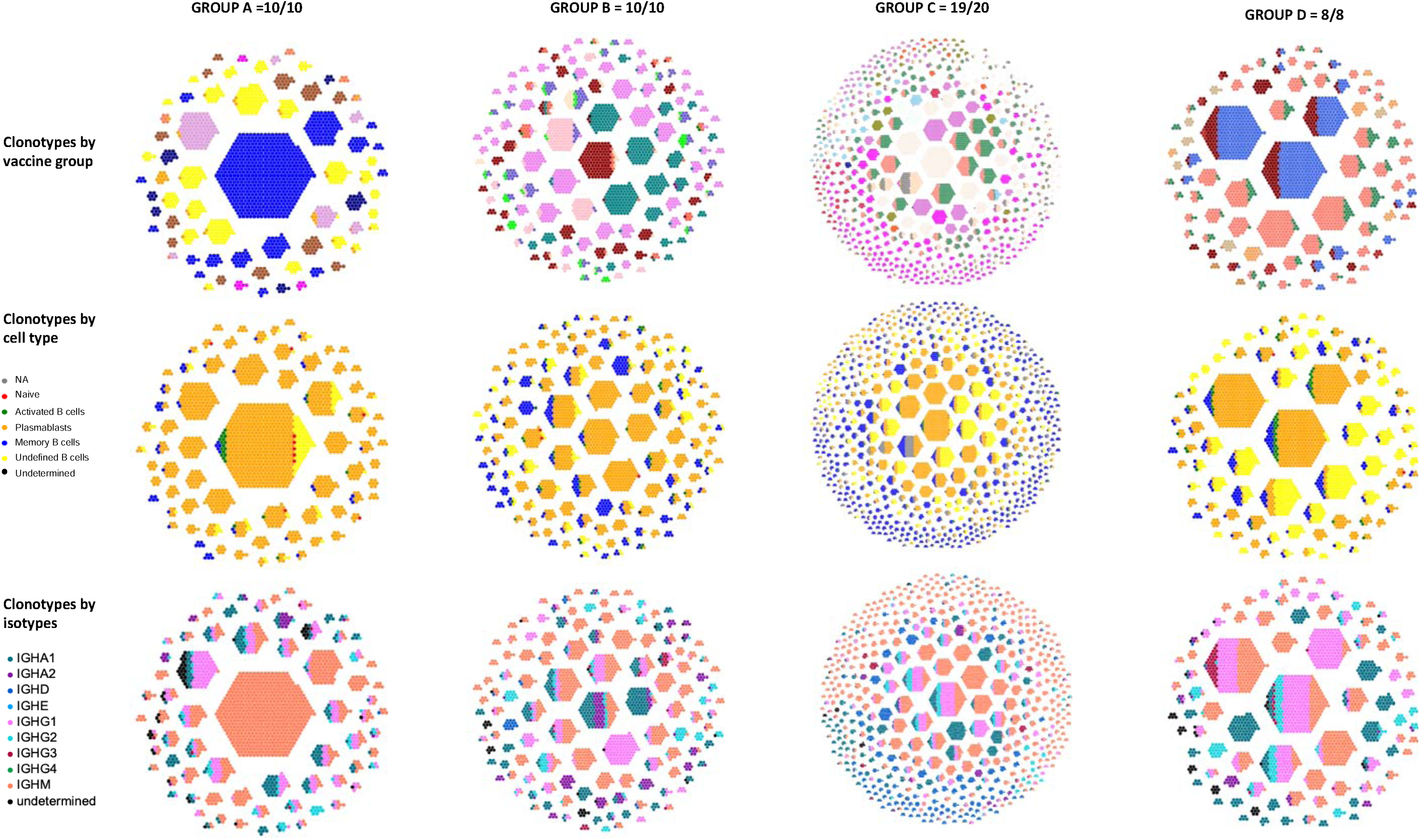
Honeycomb plots of shared clonotypes. In this figure the honeycombs are made by highlighting the vaccine groups (columns) vs number of clonotypes in total in a vaccine group (top row), type of cell (middle row) or isotypes (bottom row). Group C has the highest number of clonotypes as it had more subject compared to groups A, B and D. when loo

Within some honeycomb clonotype it is clear that different individuals from the same or different vaccine groups can share the same clonotype, Figure 6A) where two or more different individuals are within the same clonotype cluster. Similarly, honeycomb clonotypes can have different isotypes indicative of class switching (Figure 6B). Isotype distribution among the 48 samples showed, the majority of the isotypes seems represented by IgM and IgG1and IgA1, with the majority of clonotypes representing mixed IgG isotypes. Overall, IgM is the most abundant isotype for group A. Honeycomb clonotypes can also consist of different B-cell types (Figure 6C). The largest clonotypes are represented mainly by PB, while the smaller clonotypes have both MB and PB. Among all immunisation groups there were a total of 1337 clonotypes (Figure 7 - Suppl. Big table?)). Of these, 206 had between 5 to 10 cells, 61 had 11 to 20 cells, 52 between 21 to 7 and only 7 clonotypes had up to 100 cells, with 11 over 100 cells. The biggest clonotype had 834 cells, of these, the majority were filtered out as duplicates.

**Figure 7.**
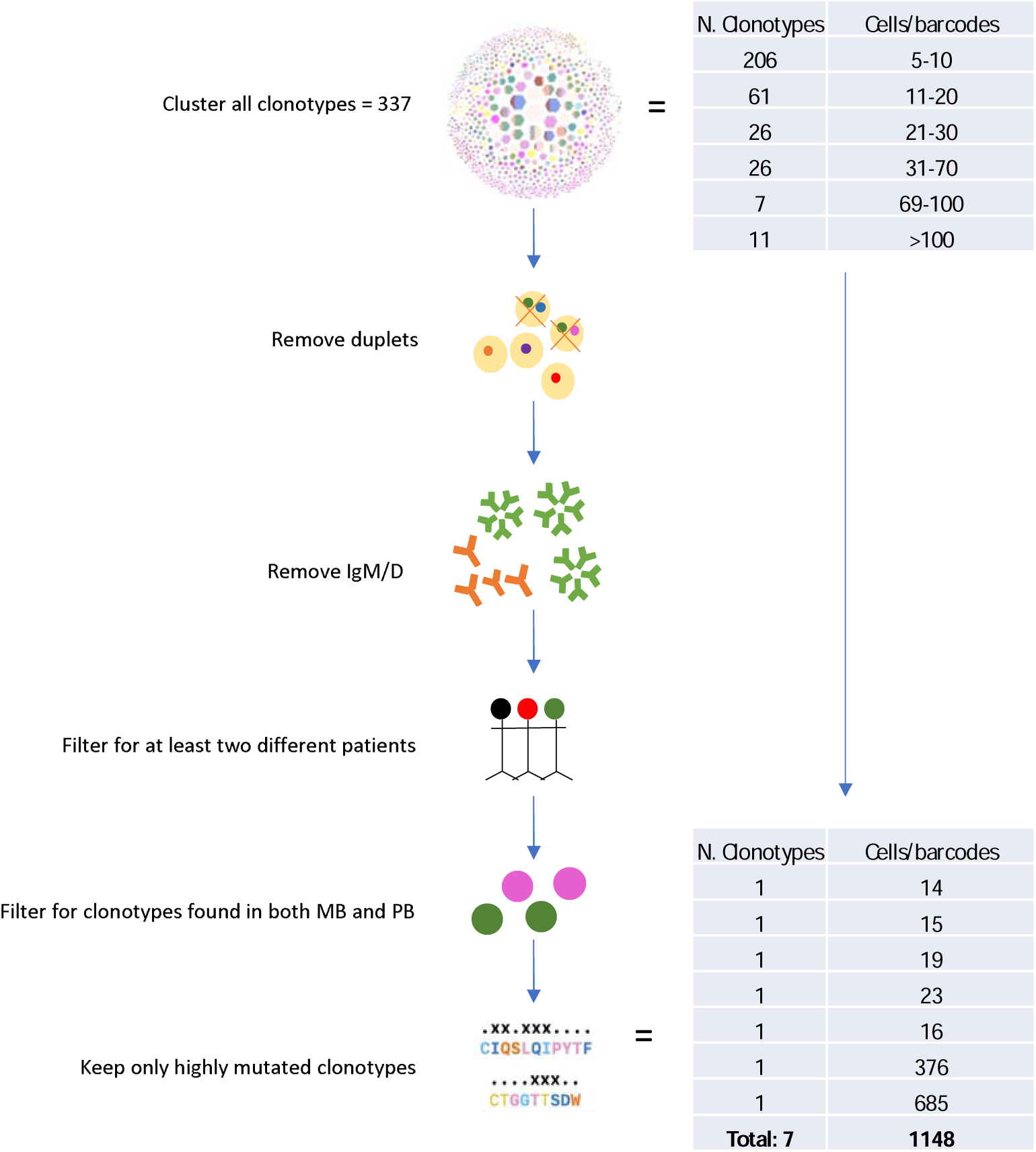
Schematic representation of the filtering process. Following data analysis, 337 clonotypes were found, divided into 6 groups and associated with a number of barcodes (cells). From these, all duplets were removed, as well as IgM/D isotypes and clonotypes were filtered to only keep those having at least two different patients, as these will be more unique than those not shared. Further filtering included for cell types (memory B cells. MB, and plasmablasts, PB) and highly mutated clonotypes. This filtering narrowed down the number to 7 clonotypes for a total of 1148 cells that we chose for further cloning experiments.

### Seven clonotypes represent functional antibodies against vaccine candidate proteins

As shown in the schematic Figure 7, we filtered down the 337 clonotypes and only kept the ones where we found different patients sharing same clonotypes, between both PB and MB. We also kept only the IgM-/IgD- and the ones with high divergence from germline. This totalled up to 7 clonotypes of which two had 376 and 685 cells and the smallest with 14 cells. In total we had 1148 cells, including the duplets. These were traced back to 10 samples we named A1-A10 to maintain the blind experiment as shown in Table 2. The table shows we only have 1 cell for samples A1, A7 and A8, 4 cells for A3, 28 cells for A2, 241 cells for A4, 87 cells for A5, 26 cells for A6, 5 cells for A9 and 14 cells for A10. This meant we removed 740 cells as duplets from further analysis. The 408 cells are the ones we believe express functional antibodies against HIV. The majority of these were either PB, MB or undefined B cells, while we found only one being a naïve cell (sample A9). The cells were mainly from two of the four vaccine groups we tested, C and D, namely ConS UFO and EDC ConS UFO. None of the cells came from groups A or B except for one barcode which belonged to group B, EDC ConM SOSIP and it was an undefined B cell.

**Table 1.** Sample vaccine regime. Only one time point, representing PBMC collected 7 days following first boost, was analyzed in this study. Patients 27/29 did no participate

| Groups | Time point 1 |
| --- | --- |
| A | ConM SOSIP = 100 µg |
| B | EDC ConM SOSIP = 100 µg |
| C | ConS UFO = 100 µg |
| D | EDC ConS UFO = 100 µg |
| E | ConS UFO = 100 µg |

**Table 2.** Summary of final cell types, vaccine groups and barcode (cell) count found in the clonotypes of interest.

| Barcode count | Cell type | Samples | Vaccine group |
| --- | --- | --- | --- |
| 1 | TB | A1 | B |
| 28 | MB/PB | A2 | C |
| 4 | PB | A3 | C |
| 241 | MB/PB/TB | A4 | D |
| 87 | MB/PB/TB | A5 | D |
| 26 | MB/PB/TB | A6 | D |
| 1 | PB | A7 | D |
| 1 | PB | A8 | C |
| 5 | MB/N | A9 | C |
| 14 | MB/PB | A10 | C |
| 408 |  |  |  |

### Phylogenetic analysis of HIV-related antibodies

We made a list of 7 clonotypes (Figure 8A-G) for further analysis via phylogeny to look into the relationships. To make the phylogenetic trees we excluded the duplets. Trees were made one for each of the 7 clonotypes selected and from these we chose the highly diversified branches for further cloning. Namely, we chose up to 3 antibodies from each tree and we matched their sequences with historical data for B cell repertoire, to exclude any duplicates antibodies which we might have found being not HIV-specific (vaccine induced). These were chosen for cloning studies currently being undertaken (data not shown).

**Figure 8A-G.**
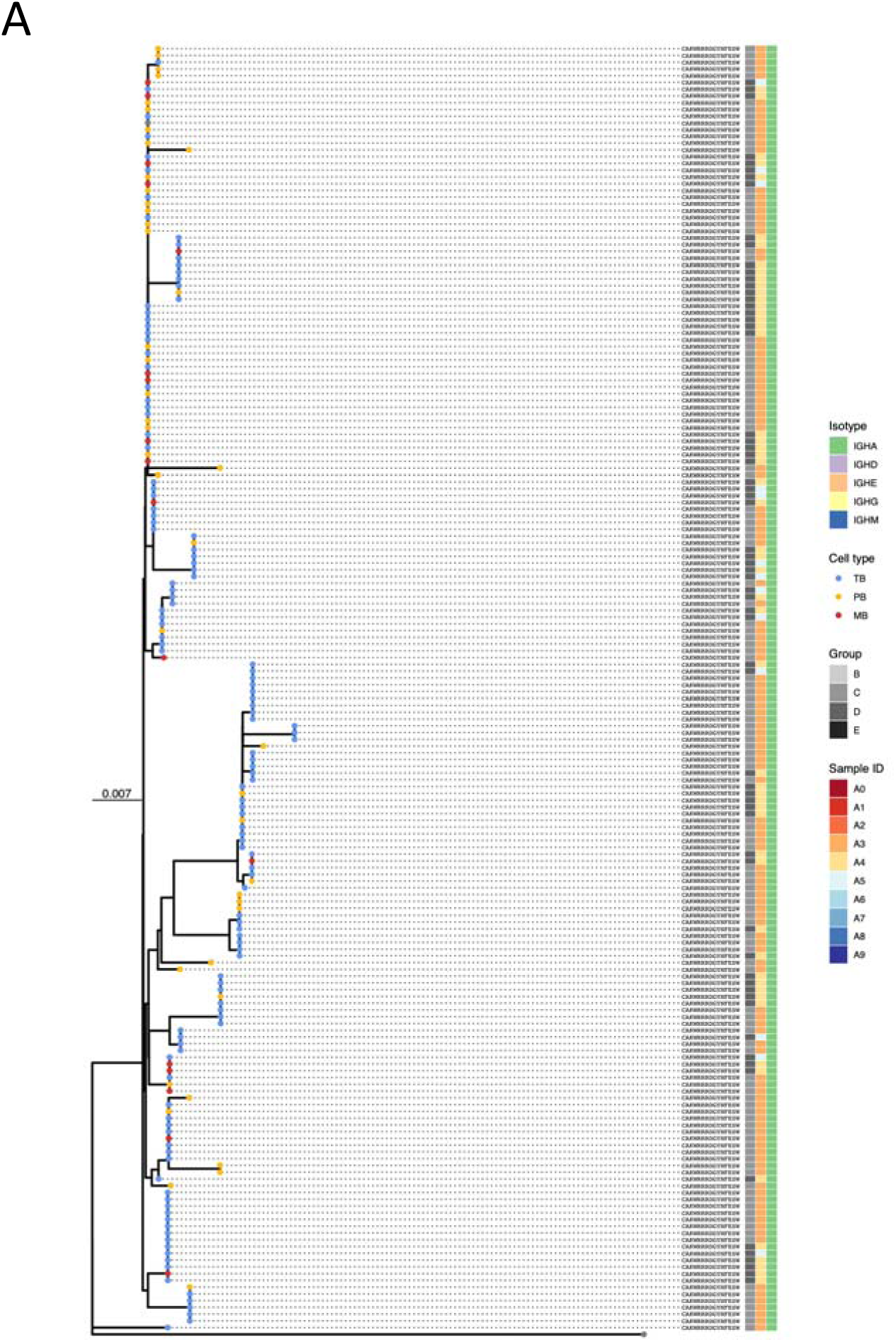
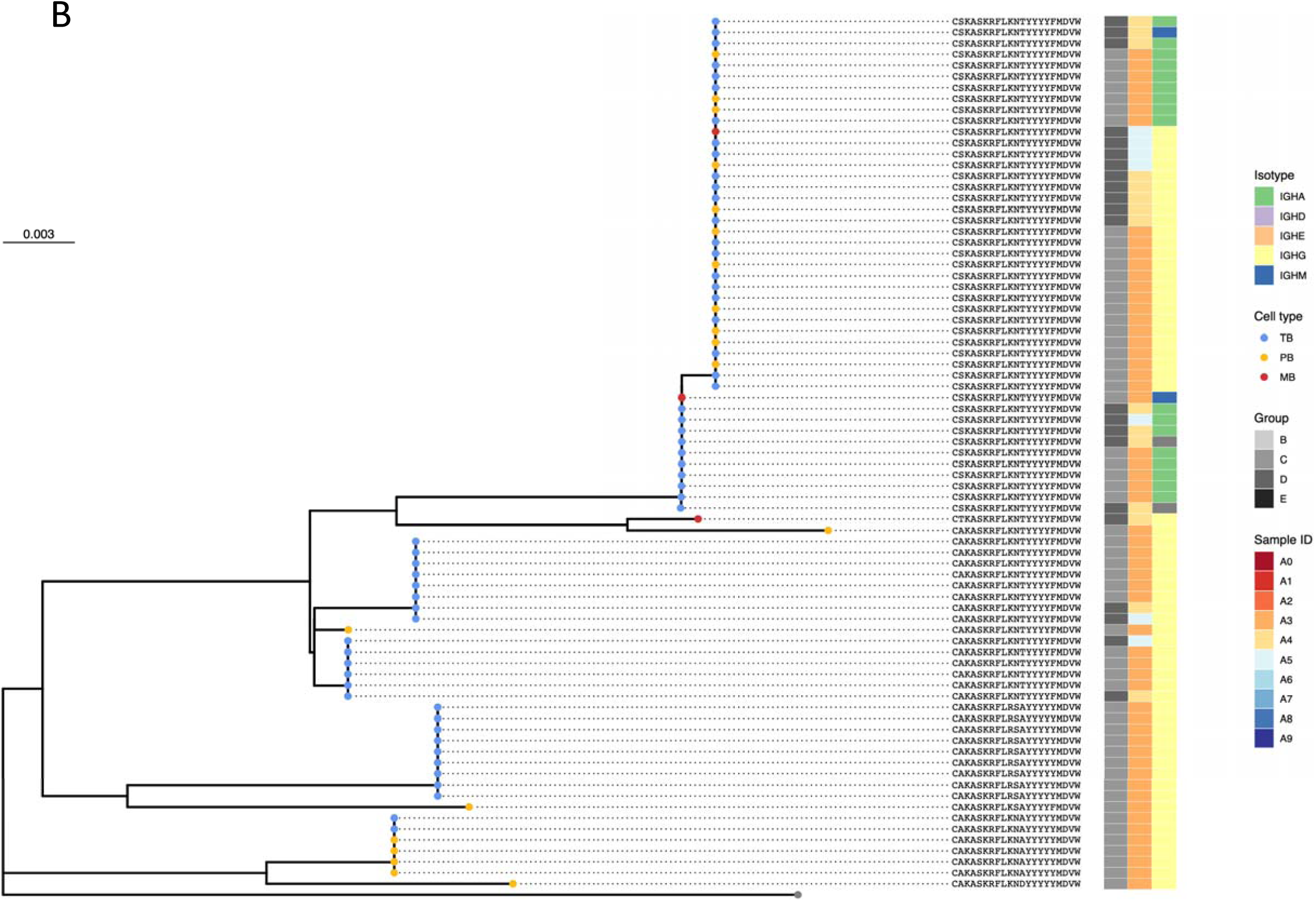
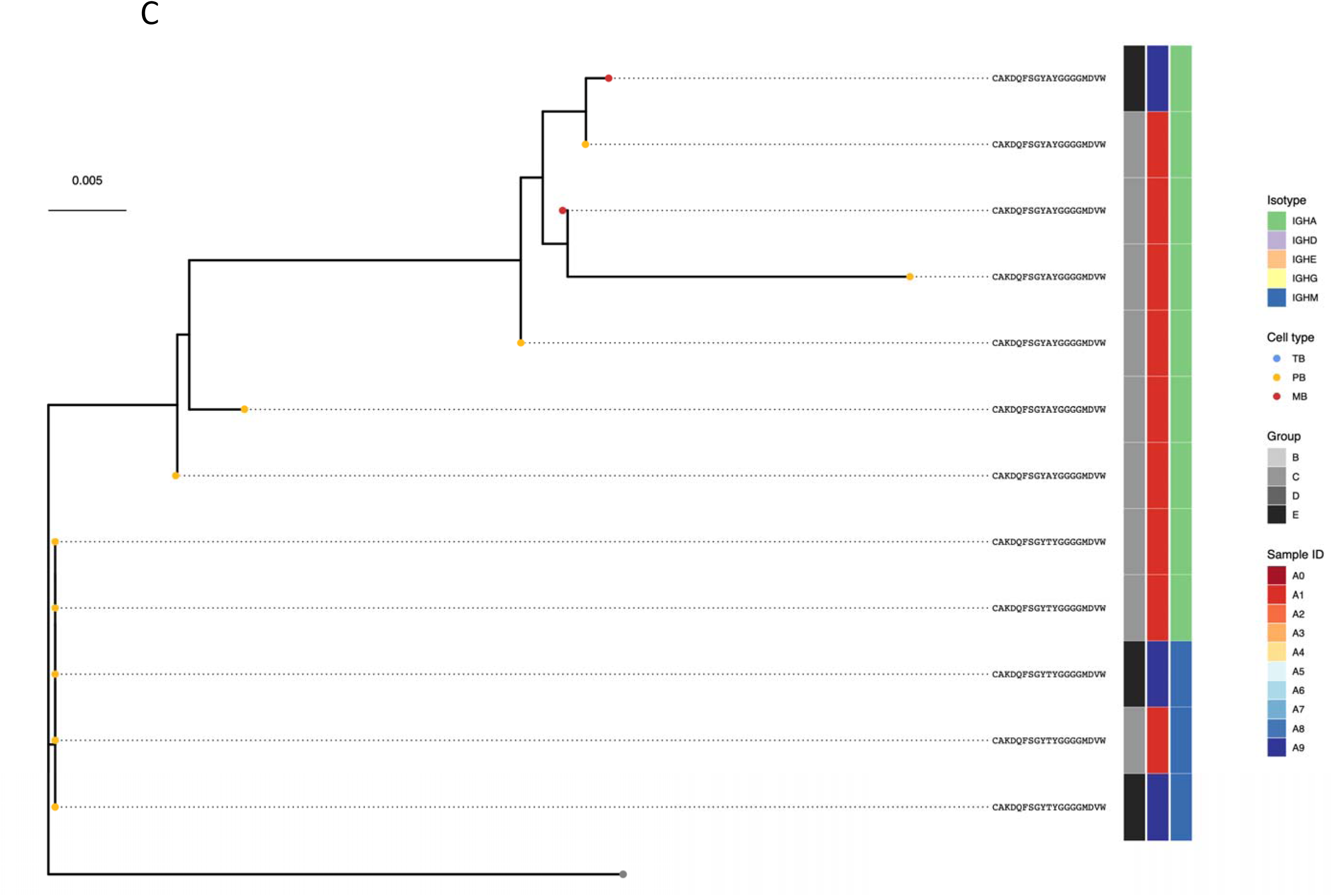
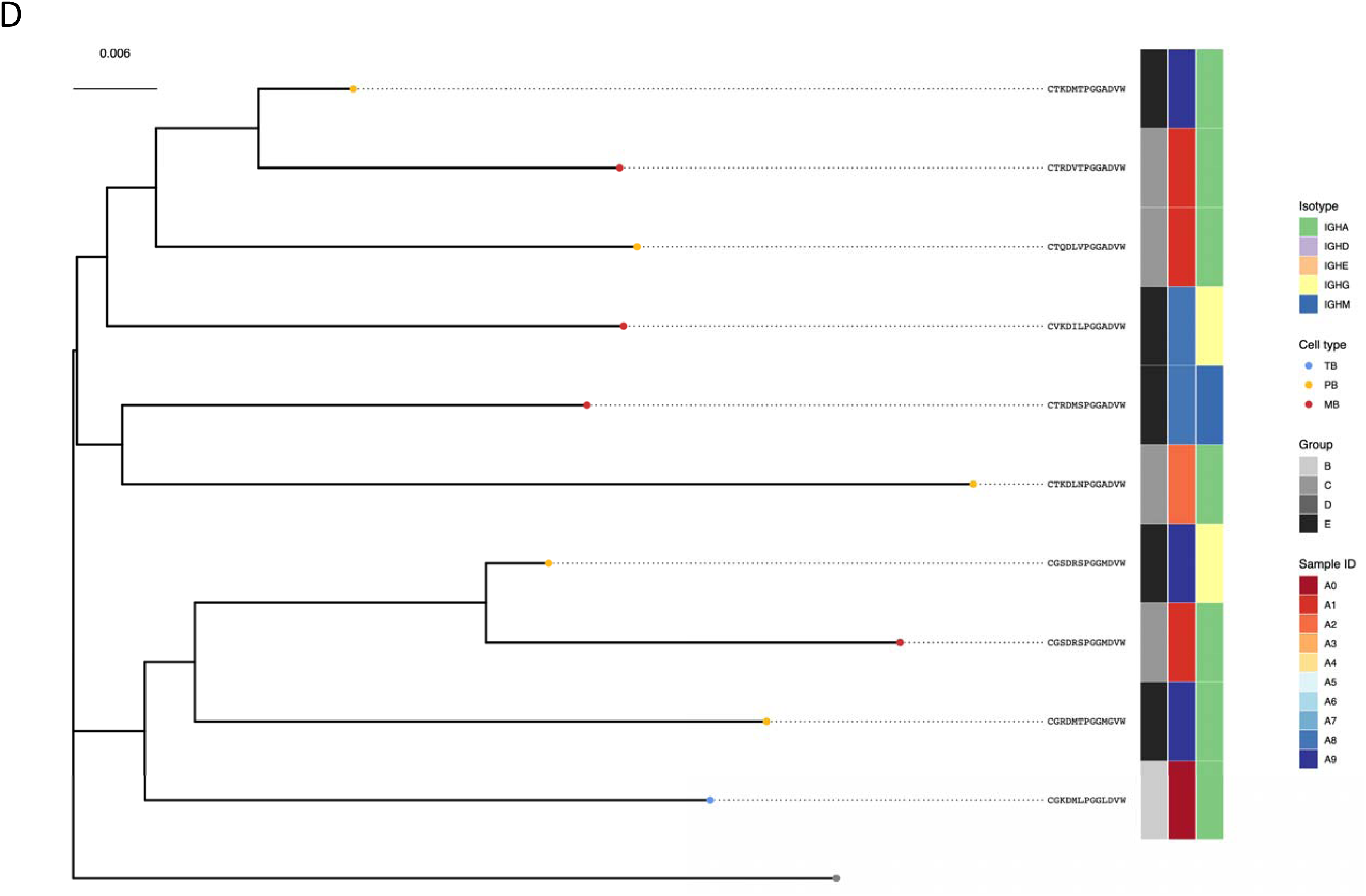
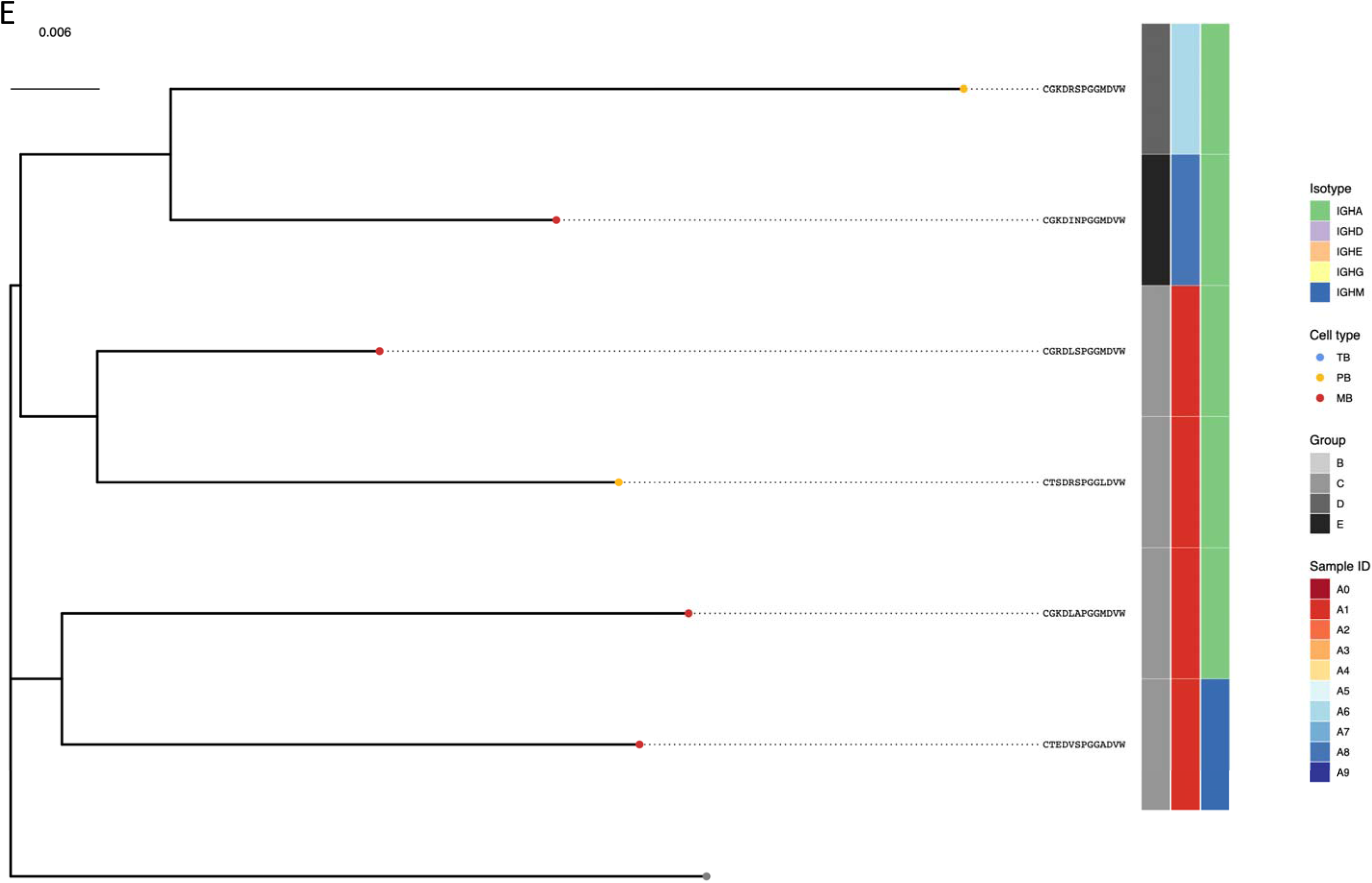
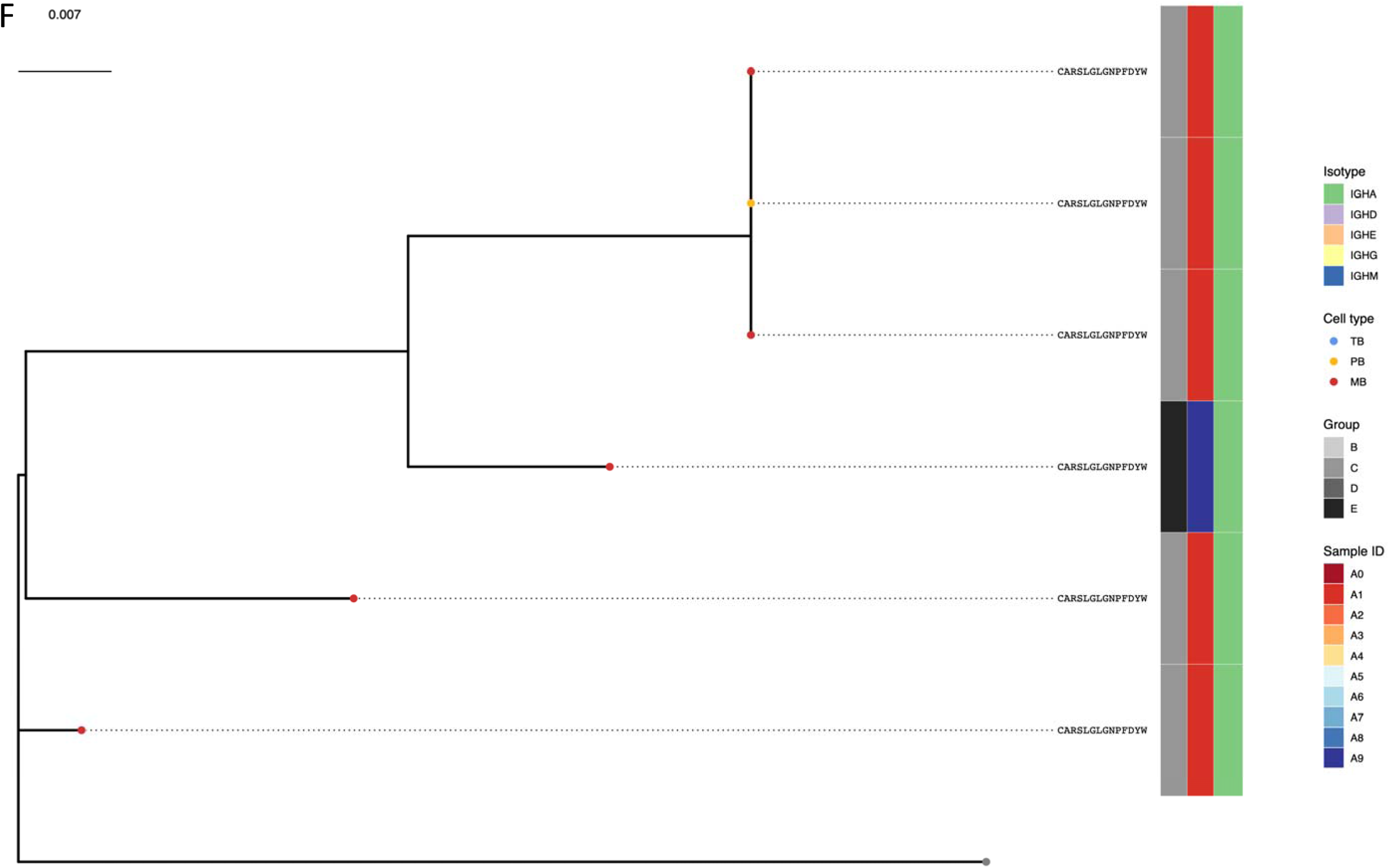
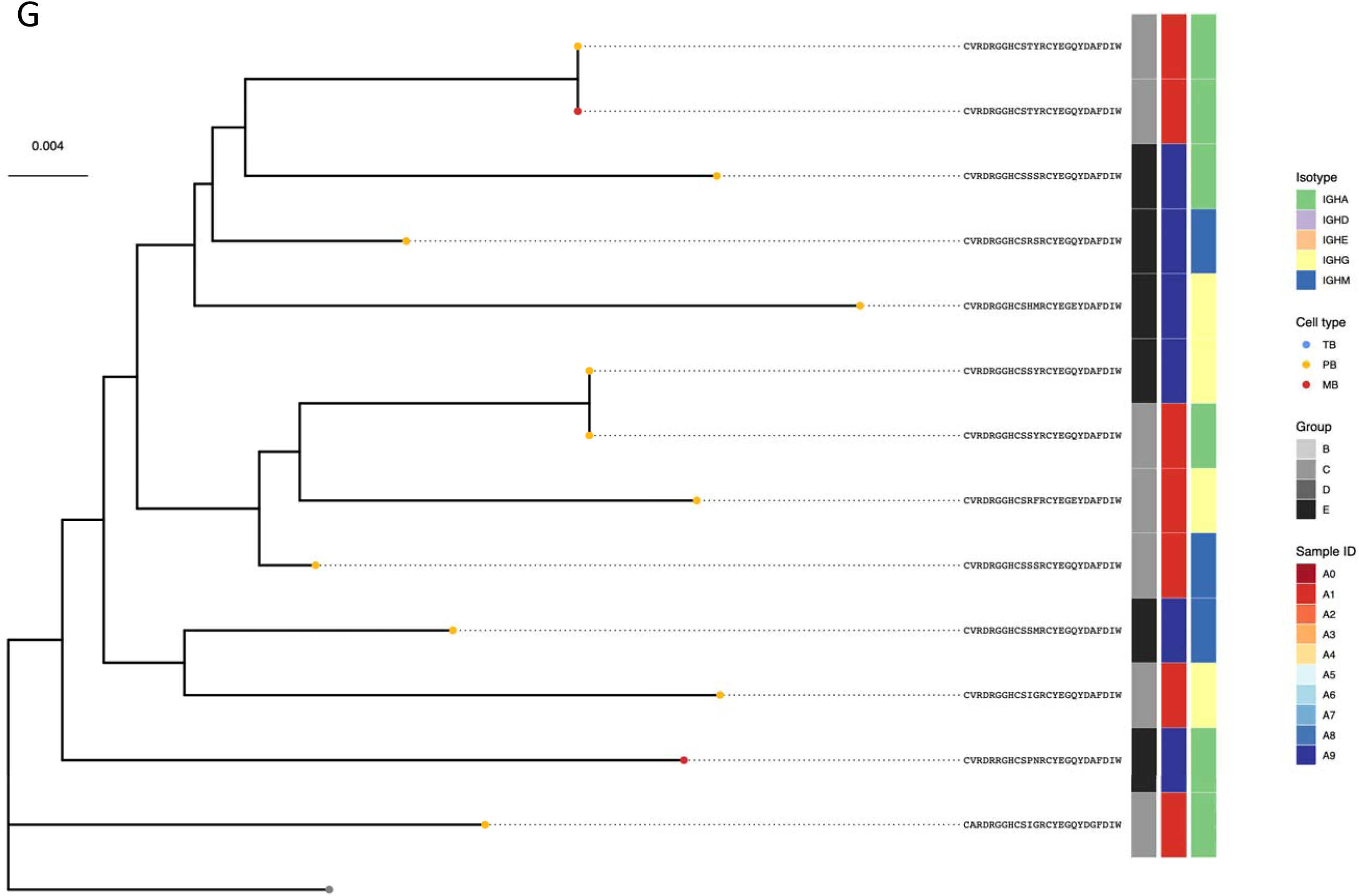
Phylogenetic trees for each for the clonotypes from Figure 7.

## Discussion

B cells are an important component of the immune response against infection. In this study we analysed the antibody response to two different vaccine candidates against HIV, ConM and ConS, and we investigate the phylogenetic relationship between the antibodies produced by the different B cell subpopulations. The novelty of the study resides in that we used single cell sequencing, by applying a multiplexing protocol which together allowed us to have a broad view of the antibody response among 48 different subjects. We used different pipelines to confirm our findings, such as those offered by Seurat [24] and 10x Genomics software. For the first time we were able to report the antibody response to two HIV-proteins being investigated in the EAVI2020 clinical trials.

Our findings highlight that among the four B cell populations, memory B cells are overrepresented, followed by plasmablasts. Notably, these two populations always clustered separately in all our samples, suggesting the diverse nature of the population. Activated B cells and naïve B cells, which instead did not cluster as much as memory B cells and plasmablasts, were also retrieved as a mixed population in memory B cells and/or plasmablasts. This is consistent with previous published data where, as infection progresses (or in our case, vaccine-induced immune response), we can see a shift of the ratios between the various B cell populations: in our experiment we did not follow the course of the infection, but by reading the cell type profile at 7 days post vaccination, where we would expect a high number of memory B cells, we found that indeed plasmablasts numbers were lower than memory B cells, as well as that at this time activated B cells can originate from memory B cells or from naïve B cells as suggested in [26]. Although we did not follow the dynamics of B cell proliferation, we were able to retrieve more populations than those manually isolated.

We based our cell type clustering for the downstream analysis on a list of internal markers which we could use with confidence to separate the various B populations (Supplementary table 1). Our initial magnetic separation was based on a negative separation where PBMCs were incubated with antibodies which separated B cells from the rest of the cells (Pan B kit from Mylteni). We then incubated the eluted B cells with CD27 antibody to further enrich our population for memory B cells and plasmablasts. We did not expect this magnetic separation to be 100% pure, since based on our downstream analysis we found both T cells (the majority CD8+ cytotoxic cells, data not shown), as well as other B cells populations, such as naïve B cells and activated B cells. Given that these latter two populations were constantly present, we included them in our analysis. Downstream analysis clearly cannot be based on the cell surface markers we used to separate the initial populations from PBMCs, since protein expression levels do not necessarily match mRNA levels. Since the data we had comes from mRNA, we had to use internal markers known to be specific for the populations of interest. We therefore built up this list based on our 10x runs (over 50 samples) as well as from the literature. Our data shows that the separation of the various B cell sub populations is consistent with the data presented in [26]. In fact, seven days post vaccine, three transcription factors are used to identify the plasmablasts populations, namely IRF4 and IRF8. As suggested in the paper, and confirmed in our analysis, IRF4 expression is higher in plasmablasts compared to naïve or activated B cells, while IRF8 has higher expression in memory B cells, naïve and activated B cells compared to plasmablasts. This suggested to us that the B cell populations isolated from total PBMCs contained not just memory B cells and plasmablasts, but also naïve and activated B cells. This was consistent among the different vaccine regimes as outlined in our experimental plan. In addition, still consistent with the literature, we found that markers such as PRDM1 and XBP1 could be used with confidence to separate plasmablasts from the rest of the populations [26], while TCL1A could be used for naive B cells separation.

When overlapping the clonotype data generated by cellranger vdj pipeline, plasmablasts had the highest numbers of clonotypes, with some clonotypes being overrepresented more than others. Based on our analysis, in some cases cell lysis that occurred before running the 10x might have generated fragments that could have ended up being captured in background GEMs. These were eliminated from our analysis. However, the majority of the samples did not undergo lysis based on the mitochondrial gene content (Supplementary Figure 2) and any cells with >5% content of MT genes was discarded. Therefore, the high number of the same clonotypes being present in a population can be genuinely assigned to that cluster, in this case plasmablasts. We generated honeycomb plots for all clonotype that passed the filtering steps, and we showed that for each vaccine group, there are patients that have bigger clonotypes that others, but also share same clonotypes. While many clonotypes are unique to individuals, shared (’public’) clonotypes are observed in different populations. We found many clonotypes that were shared between more than one patient, suggesting a strong response to the vaccine. Based on the final table we produced, the majority of the antibodies within the clonotypes and shared between the patients belong to groups C and D, and only one clonotype with a patient from group B. This suggests that ConS is more likely to induce a better antibody response compared to ConM, which we only found in one shared clonotype. In addition, we also noted that among the top most used CDR3 regions, both group C and D share the same one, namely CAKWRRRDGYNFESW. Interestingly, we also found in our final clonotype analysis a group of antibodies with this CDR3.

It is arguable that these antibodies might not be HIV-specific since we didn’t sequence the same patients before beginning the trial to investigate whether these were pre-existing clonotypes. Sorting Ag-specific cells by FACS would have been the best approach, but in our trialling of this method we found that sorting cells before running the 10x Chromium did not allow us time to successfully perform the encapsulation protocols, when considering that each sample would need to be sorted individually before hashing and loading into the 10x Chromium device, leading to an increase in cell lysis events. While this would be feasible for one or two samples, processing 12 samples for a single 10x run was not a good approach. In addition, the number of Ag-specific B cells sorted would have been very low, especially considering the cell losses during the hashing, and so even if multiple samples were combined into a single 10x run there would have still been insufficient numbers for an optimal encapsulation and thus sequencing depth. Therefore, in the absence of pre-vaccine B cells for these individuals, we used historical data from publicly available B cell repertoires. This is a common procedure when lacking baseline data as suggested in [27] where the B cell response is investigated in SARS-CoV2 patients, but for whom pre-pandemic samples are not available for the same patients.

In the last couple of years, the technologies and methodologies of single cell isolation and encapsulation have improved, for example the use of hashed antigen probes and of ‘superloading’ of the 10x Chromium platform. This would allow future studies to identify antigen-specific B cells from scRNA-Seq data without prior sorting, and to load increased numbers of multiplexed cells (greater than the 17000 used in this study) with a much higher recovery rate. In this instance, we have been able to successfully isolate single cells for analysis of the B cell repertoire without the need of antigen-sorting the cells or using pre-vaccine samples. We relied on the robustness of the bioinformatic analysis to selectively remove duplets, cells with known Antibodies against other pathogens and on phylogeny studies to make a list of possible antibodies to study further binding and neutralization assays.

| **Contributor Role** | **Role Definition** |
| --- | --- |
| **Conceptualisation** | Ideas: formulation or evolution of overarching research goals and aims: IB, PK, RS |
| **Methodology** | Development or design of methodology; creation of models: IB, CP |
| **Software** | Programming, software development; designing computer programs; implementation of the computer code and supporting algorithms; testing of existing code components: IB, SW, CM |
| **Validation** | Verification, whether as a part of the activity or separate, of the overall replication/reproducibility of results/experiments and other research outputs: IB, CP |
| **Formal Analysis** | Application of statistical, mathematical, computational, or other formal techniques to analyse or synthesise study data: IB, SW, CM |
| **Investigation** | Conducting a research and investigation process, specifically performing the experiments, or data/evidence collection: IB, CP, HC, KP |
| **Resources** | Provision of study materials, reagents, materials, patients, laboratory samples, animals, instrumentation, computing resources, or other analysis tools: PK, RS |
| **Data Curation** | Management activities to annotate (produce metadata), scrub data and maintain research data (including software code, where it is necessary for interpreting the data itself) for initial use and later reuse: IB |
| **Writing – Original Draft Preparation** | Creation and/or presentation of the published work, specifically writing the initial draft (including substantive translation): IB |
| **Writing – Review and Editing** | Preparation, creation and/or presentation of the published work by those from the original research group, specifically critical review, commentary or revision – including pre- or post-publication stages: All authors |
| **Visualisation** | Preparation, creation and/or presentation of the published work, specifically visualisation/data presentation: IB, CM, SW |
| **Supervision** | Oversight and leadership responsibility for the research activity planning and execution, including mentorship external to the core team: PK, RS |
| **Project Administration** | Management and coordination responsibility for the research activity planning and execution: PK |
| **Funding** | Acquisition of the financial support for the project leading to this publication: This work was supported by the EAVI2020 grant 681137 to RS and PK. |

## Data Availability

Data will be available upon reasonable request

## Acknowledgements

The authors thank Dr Hamish King, Walter and Eliza Hall Institute of Medical Research for useful comments and advice.

