## Supplementary figures and images for "B cell receptor analysis using single cell sequencing reveals preferential antibody expression in a HIV candidate vaccine study"

### Supplementary figures

#### Slide 1
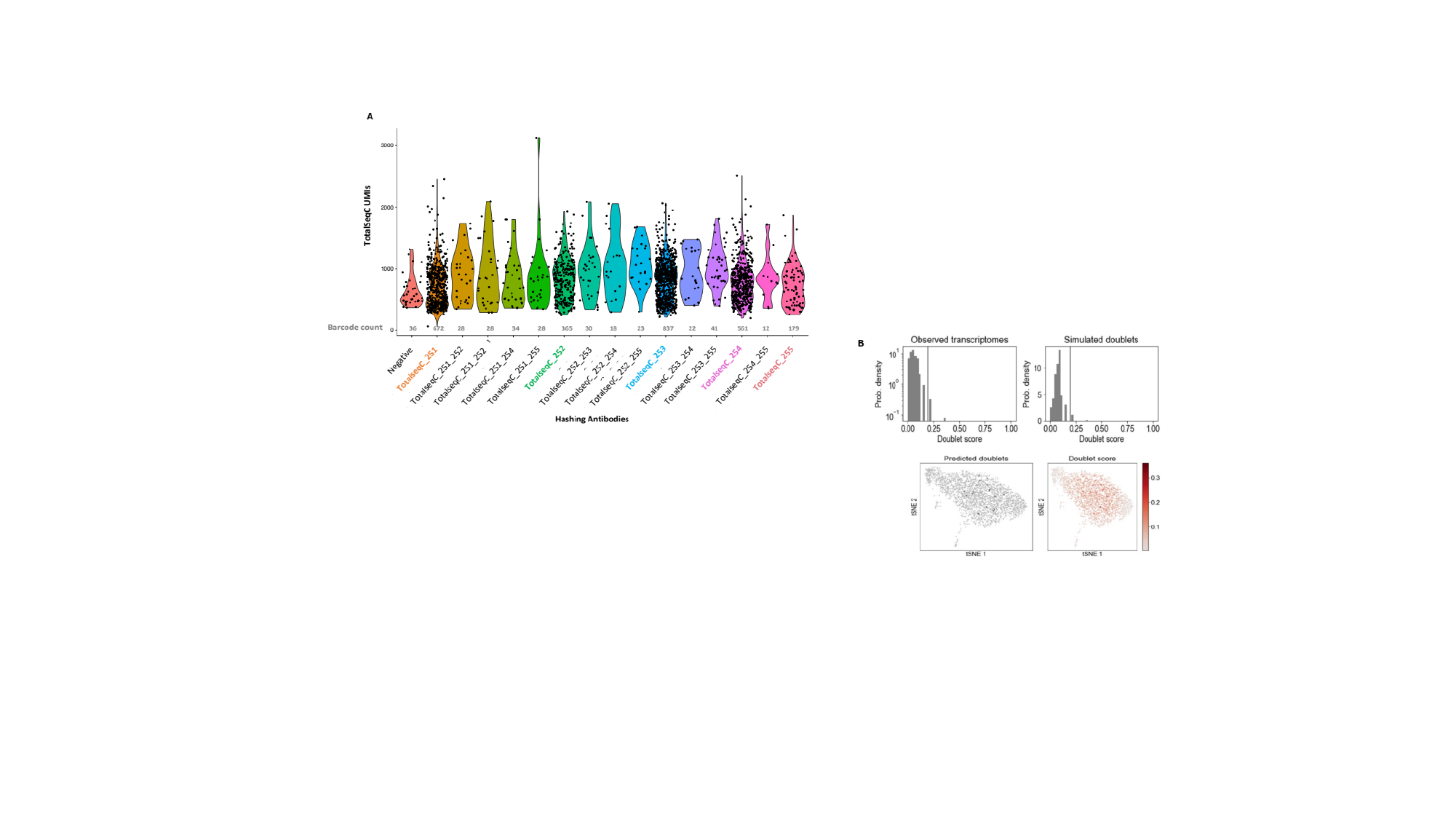

#### Slide 2
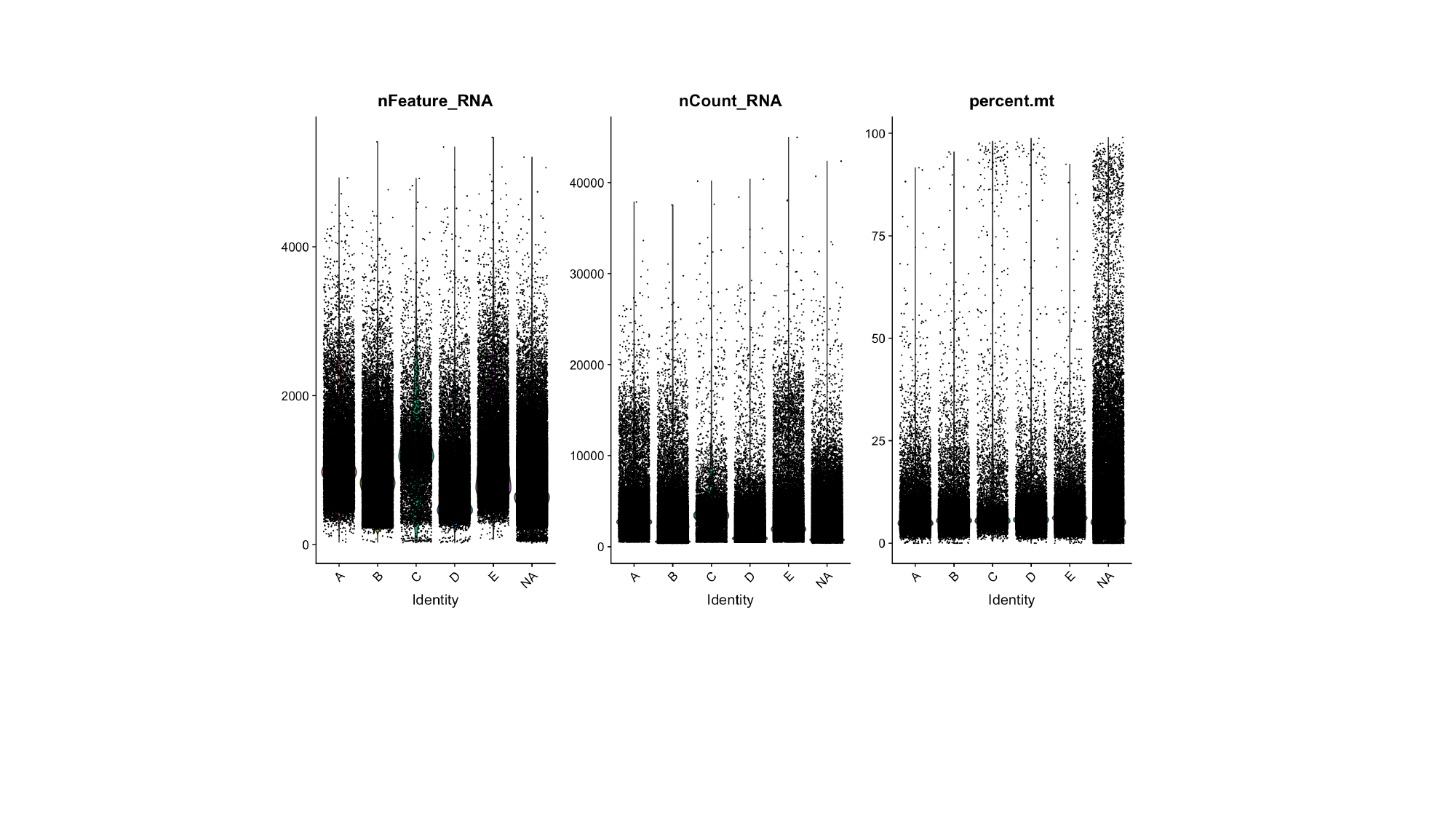

#### Slide 3
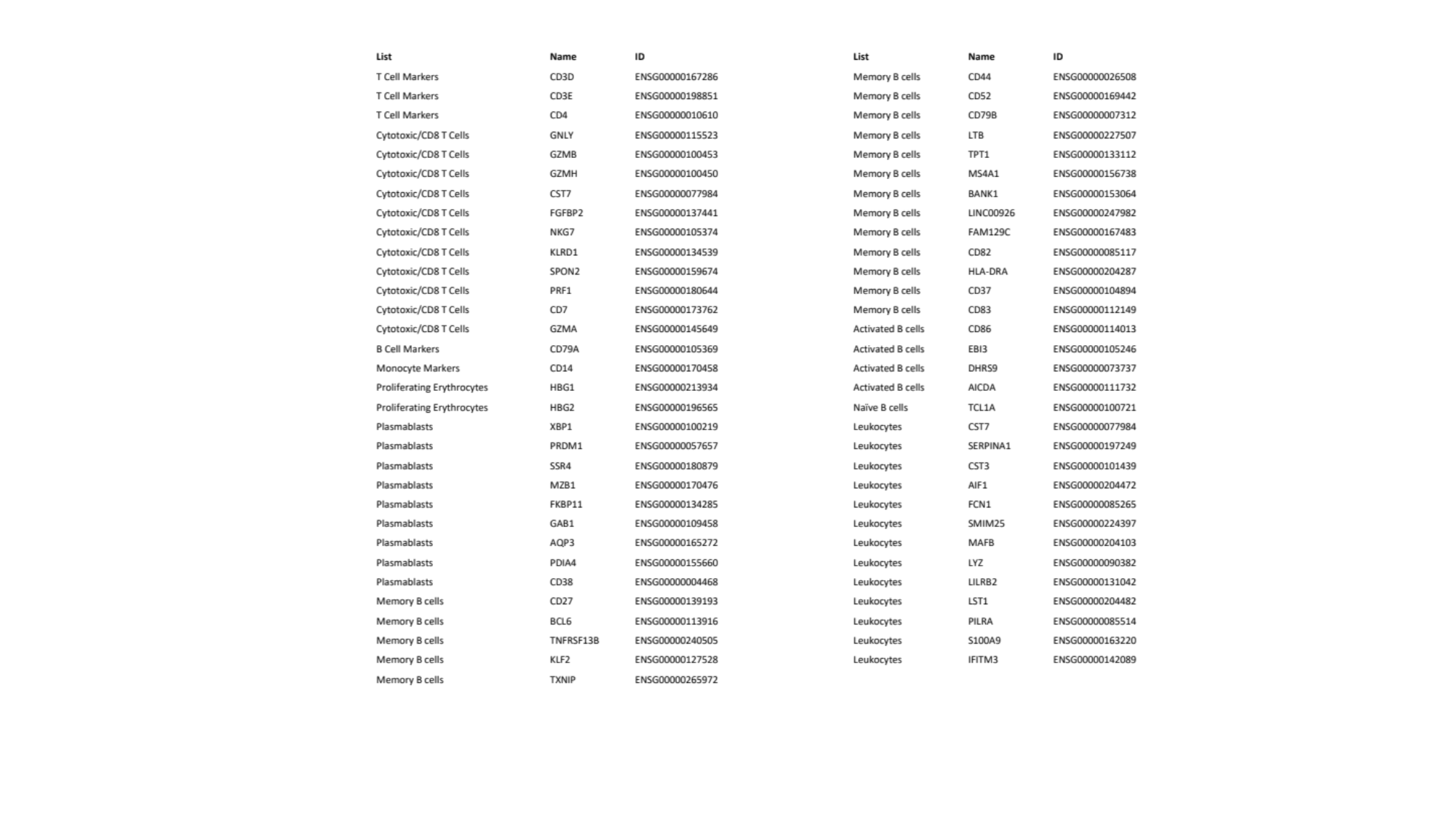
